# CD19^+^IgD^+^CD27^-^ naïve B Cells as predictors of humoral response to COVID-19 mRNA vaccination in immunocompromised patients

**DOI:** 10.1101/2021.08.11.21261898

**Authors:** Eduard Schulz, Isabel Hodl, Patrick Forstner, Stefan Hatzl, Nazanin Sareban, Martina Moritz, Johannes Fessler, Barbara Dreo, Barbara Uhl, Claudia Url, Andrea Grisold, Michael Khalil, Barbara Kleinhappl, Christian Enzinger, Martin H. Stradner, Hildegard Greinix, Peter Schlenke, Ivo Steinmetz

## Abstract

Immunocompromised patients are considered high-risk and prioritized for vaccination against COVID-19. We aimed to analyze B-cell subsets in these patients to identify potential predictors of humoral vaccination response. Patients (n=120) suffering from hematologic malignancies or other causes of immunodeficiency and healthy controls (n=79) received a full vaccination series with an mRNA vaccine. B-cell subsets were analyzed prior to vaccination. Two independent anti-SARS-CoV-2 immunoassays targeting the receptor-binding domain (RBD) or trimeric S protein (TSP) were performed three to four weeks after the second vaccination. Seroconversion occurred in 100% of healthy controls, in contrast to 67% (RBD) and 82% (TSP) of immunocompromised patients, while only 32% (RBD) and 22% (TSP) achieved antibody levels comparable to those of healthy controls. The number of circulating CD19^+^IgD^+^CD27^-^ naïve B cells was strongly associated with antibody levels (ρ=0.761, P<0.001) and the only independent predictor for achieving antibody levels comparable to healthy controls (OR 1.07 per 10-µl increase, 95%CI 1.02–1.12, P=0.009). Receiver operating characteristic analysis identified a cut-off at ≥61 naïve B cells per µl to discriminate between patients with and without an optimal antibody response. Consequently, measuring naïve B cells in immunocompromised hematologic patients could be useful in predicting their humoral vaccination response.

## INTRODUCTION

Coronavirus disease 2019 (COVID-19) results in increased morbidity and mortality in immunocompromised patients.(1-3) Immunodeficiency can be primary (PID) due to underlying genetic causes such as common variable immunodeficiency or secondary (SID) resulting from hematologic malignancies (HM), immunosuppressive therapies, or hematopoietic stem cell transplantation (HSCT). In a recent study of 100 patients with COVID-19 disease, patients with PID and SID demonstrated higher morbidity and mortality than the general population, while the outcomes of individuals suffering from SID were the worst.(1) In patients with HM and COVID-19, a mortality rate of 34% (95% confidence interval [CI]: 28–39) has been reported in adults in a recent meta-analysis including 3377 predominantly hospitalized patients from 3 continents.(4) Interestingly, patients on systemic anticancer treatment had a similar risk of death compared to patients without therapy (RR 1.17, 95% CI: 0.83-1.64). Risk of death was highest in patients with acquired bone marrow failure syndromes (53%, 95% CI: 34-72), followed by acute leukemias (41%, 95% CI: 30-52), myeloproliferative neoplasms (34%, 95% CI: 19-51), plasma cell dyscrasias (33%, 95% CI: 25-41), lymphomas (32%, 95% CI: 18-48), and chronic lymphocytic leukemias (CLL) (31%, 95% CI: 23-40), respectively.

Patients with HM can be immunocompromised due to the underlying malignancy itself, prior or ongoing treatments with a high degree of immunosuppressive effects such as corticosteroids, B-cell depleting therapies, HSCT and other cellular therapies. In individuals with these risk factors, lower rates of seroconversion have been reported after COVID-19 infection whereas other cancer patients developed antibody response similar to healthy individuals.(5, 6) Roeker and colleagues observed that 67% of patients with CLL developed IgG antibodies to SARS-CoV-2 nucleocapsid and the seroconversion rate among recipients of HSCT and chimeric antigen receptor (CAR) T-cell therapy was similar at 66%.(7, 8)

Due to the high risk of severe COVID-19 in immunocompromised patients, they are considered a high priority for COVID-19 vaccination.(9-13) However, trials of the currently approved COVID-19 vaccines have excluded individuals diagnosed with immunodeficiency or malignancy; therefore, information on the efficacy and safety of the vaccines in these patients is sparse.(14-17) It is well known that vaccinations in patients early after HSCT and anti-CD20 therapies as well as with several forms of PID have low efficacy.(18-20) The humoral immune response to a recombinant zoster vaccine in patients with B-cell lymphoma and CLL was between 20% and 50% compared to 80% in patients with other HM.(21)

Lack of antibody responses after COVID-19 vaccination and significantly lower antibody levels in responders have been reported in HM patient cohorts in general(5, 22, 23) and in selected patients with multiple myeloma, CLL, and non-Hodgkin’s lymphoma.(24-28) Low efficacy of COVID-19 vaccinations was observed when administered soon after HSCT and anti-CD20 therapies.(9, 18, 19) Furthermore, immunocompromised patients due to inborn errors of immunity or autoimmune rheumatic disease (AIRD) demonstrated also reduced rates in seroconversion, especially when given B-cell-depleting therapy and glucocorticoids.(14-17)

Peripheral B cells are needed for humoral vaccination responses.(29) However, the number of circulating B cells or of a certain B-cell subset associated with a humoral vaccination response comparable to healthy individuals is unknown. A marker predictive of vaccination response would aid to schedule vaccinations in the immunocompromised patients to achieve an optimal vaccination response.

We hypothesize that specific B-cell subsets have to be present in immunocompromised individuals to enable a humoral vaccination response. Herein, we used data from an interim analysis of the prospective, open-label, phase IV CoVVac trial (NCT04858607) to test this hypothesis.

## MATERIALS AND METHODS

### Study design and participants

We report the data of an interim analysis of the CoVVac trial (NCT04858607), which is an ongoing open-label, phase IV, prospective, monocentric study at the Medical University of Graz, Austria. After approval by the ethics committee of the Medical University of Graz in April 2021 (EK 1128/2021), patients with inborn errors of immunity, hematological malignancies, those receiving B-cell-depleting therapy, and healthy controls were recruited before receiving their first dose of COVID-19 vaccine. The detailed study protocol is provided in the Supplementary Information. In brief, blood was drawn before the first vaccination with BNT162b2 (BioNTech/Pfizer) or mRNA-1273 (Moderna) for peripheral blood mononuclear cell (PBMC) isolation and lymphocyte phenotyping. The second vaccination was administered 21 (BNT162b2) or 28 days (mRNA-1273) after the first. Blood sampling was performed 21–28 days after the second vaccination to analyze the COVID-19-specific antibody response as the primary endpoint.

### Lymphocyte phenotyping

Blood samples from the baseline visit were processed within 4 hours for analysis by flow cytometry. For lymphocyte phenotyping, ethylenediaminetetraacetic acid whole blood was stained for CD3, CD4, CD8, CD45, CD16, CD56, and CD19, as previously described.(30) For B-cell phenotyping, PBMCs were isolated from lithium heparin whole blood by Ficoll gradient density centrifugation. One million PBMCs were incubated with the following antibodies: CD19-VioGreen, anti-IgD-VioBlue, CD24-PerCP-Vio700, CD38-FITC, CD27-APC, CD86-PE-Vio770, CD21-APC-Vio770, and anti-IgM-PE (Miltenyi Biotec, Bergisch Gladbach, Germany). Samples were measured using a FACSLyric flow cytometer (BD Biosciences, Franklin Lakes, NJ, USA). Data were analyzed using the FACSSuite (BD Biosciences). The gating strategy is shown in Supplemental Figure 1.

### Antibody assays

Blood was obtained before first and 21–28 days after the second vaccination. Serum was aliquoted, frozen, and stored at -80°C until analysis was performed in batches. Two commercially available CE-certified serological tests were performed according to the manufacturers’ protocols to determine and quantify specific antibodies against SARS-CoV-2. Specific IgG was determined using the Roche Elecsys anti-SARS-CoV-2 S electrochemiluminescence immunoassay targeting the receptor-binding domain of the viral spike protein using a Cobas e 801 analytical unit (Roche Diagnostics GmbH, Mannheim, Germany).(27, 29) Its quantification range lies between 0.4 and 2500 U/ml, with a cut-off of 0.8 U/ml for positivity. Specific IgG was measured by Liaison SARS-CoV-2 TrimericS IgG test on Liaison XL (DiaSorin, Saluggia, Italy), which is a chemiluminescence immunoassay quantifying antibodies that target the trimeric S protein.(28) Results are provided in binding antibody units (BAU) with a quantification range of 4.81–2080 BAU/ml. Values ≥33.8 BAU/ml were considered positive.

### Statistical analysis

All statistical analyses were performed using Stata 16.1 (Stata Corp., Houston, TX, USA). Continuous data were reported as medians (25th–75th percentile) and categorical data as absolute frequencies (%). Correlations and associations between antibody response and other variables were examined using Spearman’s rank-based correlation coefficients, rank-sum test, and χ^2^-squared tests. R^2^-statistics were obtained from multiple linear regression models with antibody response as the dependent variable. The optimal cut-off to separate patients with and without vaccination response was assessed by employing a maximized Youden’s index within a receiver operating characteristic (ROC) analysis. Logistic models were used for univariate and multivariate modeling of the vaccination response. The Kruskal–Wallis test was used for continuous variables when comparing three or more treatment groups. The Kruskal-Wallis H test was used as a post-hoc test to determine between-group differences.

## RESULTS

### Study population

Data of 199 study participants who completed their full vaccination schedule were included in the efficacy analysis. Of these, 79 were healthy participants and 120 were immunocompromised patients. All study participants were vaccinated with mRNA-1273, with only two healthy individuals (1%) receiving BNT162b2. A total of 140 adverse events occurred after vaccination, with the most common being pain at the injection site, headache, fever, and fatigue. Four severe adverse events (hospitalization and death due to HM and bone fractures) were considered unrelated to vaccination. Participant characteristics are listed in Table 1. Diagnoses, immunosuppressive treatments, and antibody responses for subgroups are detailed in Supplementary Table 1.

**Table 1.** Baseline characteristics of the study population. Data are reported as medians [25th-75th percentile] and absolute frequencies (%). P-values are from rank-sum tests, χ2-tests, and Fisher’s exact tests, as appropriate. Significant P-values are highlighted in bold type. Abbreviations: n, number; HSCT, hematopoietic stem cell transplantation

| Variable | Healthy (n=79) | Immunocompromised (n=120) | P |
| --- | --- | --- | --- |
| Age (years) | 51 [36-56] | 58 [50-65] | **<0.001** |
| Female gender n (%) | 45 (57) | 65 (54) | 0.698 |
| Body mass index (kg/m<sup>2</sup>) | 23.7 [21.6-26.4] | 24.8 [22.7-27.8] | **0.039** |
| **Vaccine** |  |  | 0.080 |
| mRNA-1273 (Moderna) | 77 (97) | 120 (100) |  |
| BNT162b2 (BioNTech/Pfizer) | 2 (3) | 0 (0) |  |
| **Immunodeficiency group** |  |  | n/a |
| Primary immunodeficiency n (%) |  | 25 (21) |  |
| Autoimmune disease n (%) |  | 39 (32) |  |
| Hematologic disease n (%) |  | 56 (47) |  |
| **B-cell depleting therapy** |  |  | n/a |
| None n (%) |  | 44 (37) |  |
| Rituximab n (%) |  | 35 (29) |  |
| Ocrelizumab n (%) |  | 6 (5) |  |
| HSCT n (%) |  | 35 (29) |  |
| Days since B-cell depletion |  | 166 [69-545] |  |

### Immunogenicity of mRNA vaccines

Antibody responses were assessed using two different assays. All healthy controls demonstrated seroconversion with high antibody titers (Roche median: 2500 U/ml; DiaSorin median: 2080 BAU/ml). In immunocompromised patients, the seroconversion rates and antibody levels were significantly lower than those in healthy controls (Table 2 and Figure 1); 67% (n = 80) and 82% (n = 98) of patients demonstrated a humoral response with antibody levels within the quantification range of Roche and DiaSorin assays, respectively. Since the clinical significance of antibody levels close to the limit of detection was unclear, we defined more stringent thresholds for our patients, namely a “stringent response” as reaching at least the lowest antibody level of the healthy individuals from our cohort (Roche ≥1000 U/ml; DiaSorin ≥2000 BAU/ml). According to this definition, only 32% (Roche) and 22% (DiaSorin) of patients had a stringent antibody response. This difference was not statistically significant between the two tests (P = 0.108). Patients who received anti-CD20 therapy, including the majority of patients with AIRD, had the lowest rate of stringent response (≤10%). Interestingly, patients who received HSCT demonstrated a relatively high rate of stringent response [37% (13/35)]. There was no statistically significant difference between allogeneic and autologous HSCT (40% vs. 30%, respectively; P = 0.541). The antibody levels of both assays showed an excellent correlation with each other in the patient population (ρ = 0.915, P<0.001, R^2^ = 0.841; Table 2; Figure 2A), as well as in the entire study population (Supplementary Figure 2A). Since the Roche assay is more widely used in research and shows a good correlation with live virus neutralization tests in vaccinated individuals(22, 27, 29, 31), we focused on the Roche assay for subsequent analyses to ensure comparability with other studies.

**Fig 1.**
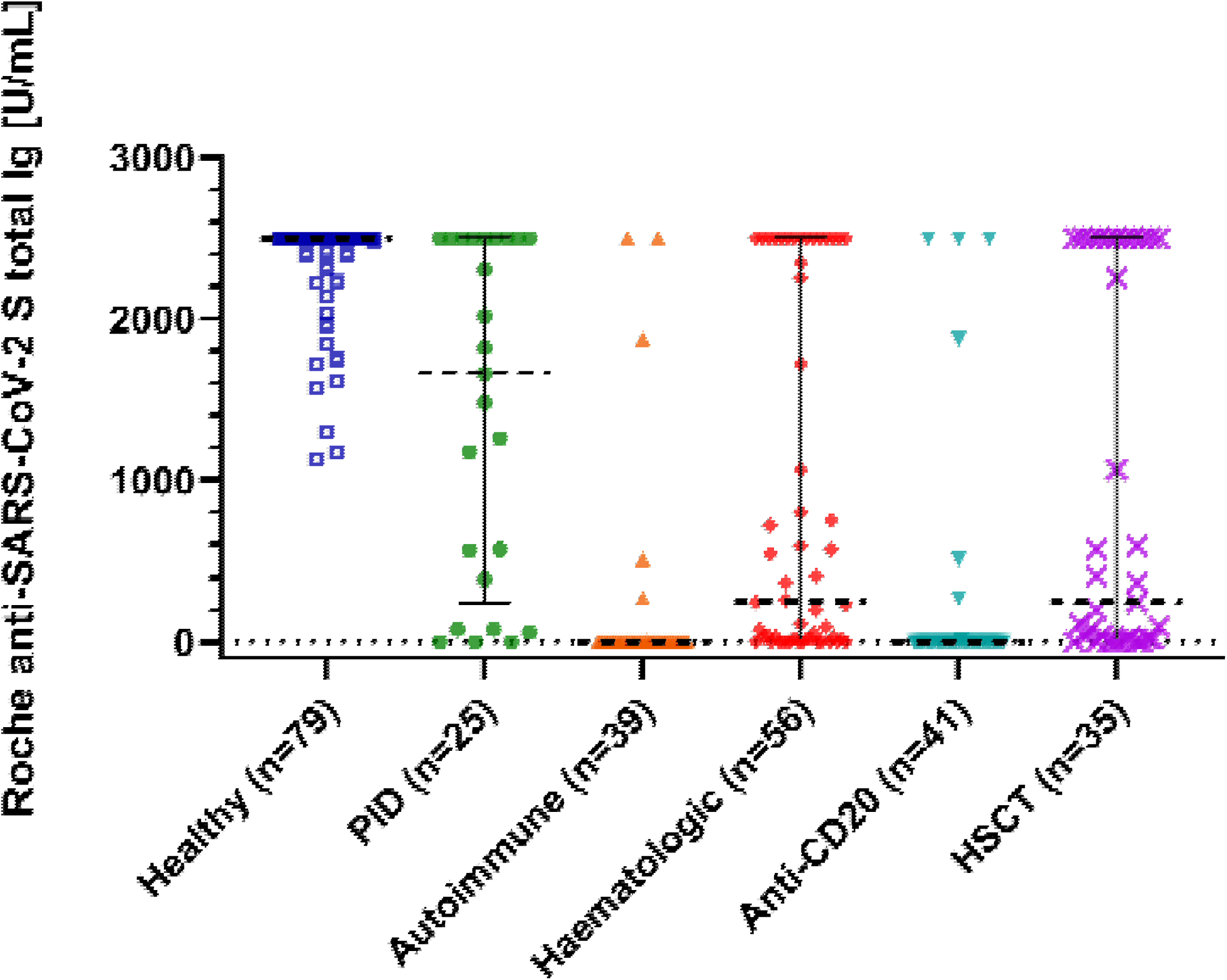
Immune response to SARS-CoV-2 mRNA vaccination. The scatter plot shows total immunoglobulin levels for healthy controls, immunodeficient patients and patients after anti-CD20 therapy or hematopoietic stem cell transplantation (HSCT). P is <0.001 between all groups calculated with the Kruskal-Wallis test and post hoc analysis. Lines are medians with interquartile range. The Plot was drawn with GraphPad Prism Version 9.2.0.332.

**Table 2.** Antibody Response to Vaccination. Data are reported as medians [25th-75th percentile] and absolute frequencies (%). P-values are from rank-sum tests, χ2-tests, and Fisher’s exact tests, as appropriate. Significant P-values are highlighted in bold type. Any response, any seroconversion; BAU, binding antibody unit; DiaSorin stringent response, DiaSorin SARS-CoV-2 TrimericS IgG ≥2000 BAU/ml; HSCT, hematopoietic stem stell transplantation; Roche stringent response, Roche anti-SARS-CoV-2 S total antibody titer ≥1000 U/ml.

|  | **Healthy (n=79)**<br>**N (%)** | **Immunocompromised (n=120) N (%)** | **P** |
| --- | --- | --- | --- |
| Roche any response | 79 (100) | 80 (67) | **<0.001** |
| Roche stringent response | 79 (100) | 38 (32) | **<0.001** |
| Roche U/ml | 2500 [2500-2500] | 67 [0-1947] | **<0.001** |
| DiaSorin any response | 79 (100) | 98 (82) | **<0.001** |
| DiaSorin stringent response | 79 (100) | 26 (22) | **<0.001** |
| DiaSorin BAU/ml | 2080 [2080-2080] | 233 [12-1760] | **<0.001** |

**Fig 2.**
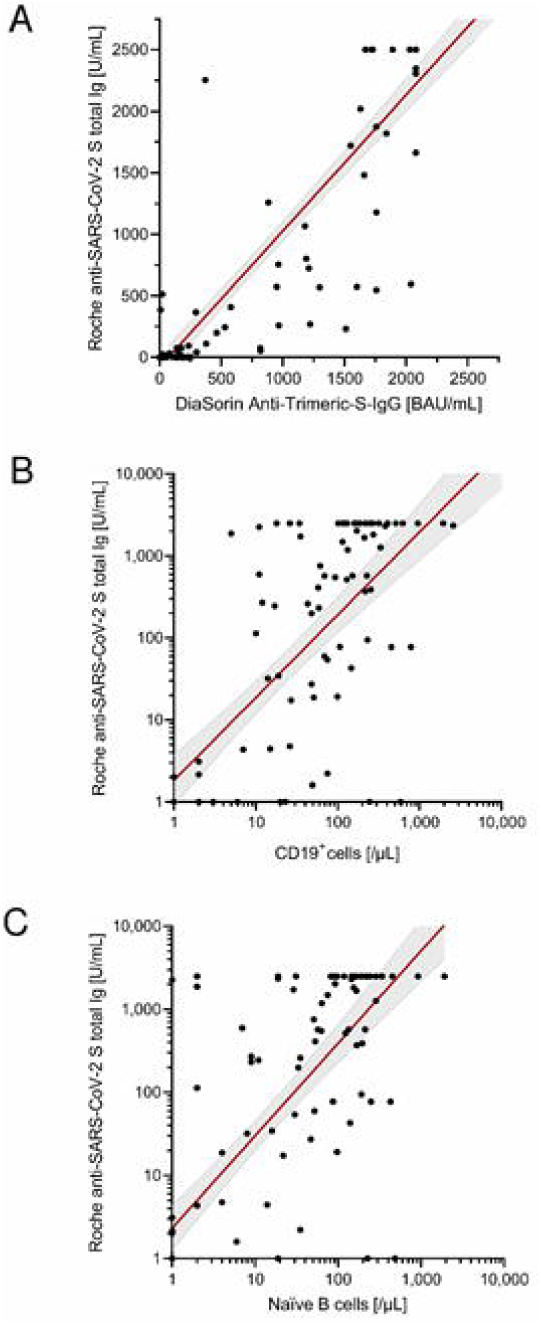
Correlation of antibody levels determined by Roche anti-SARS-CoV-2 S assay. (A) DiaSorin SARS-CoV-2 TrimericS IgG. (B) Absolute number of B cells (CD19+ cells). (C) Absolute number of naïve B cells. Scatter plots indicate a linear regression line including a 95% confidence interval. In case of A and B, regression line corresponds to transformed data using x=log((x+1) and y=log((y+1)), respectively.

### Correlation of antibody levels with the amount of B-cell subsets

The total number of B cells and all B-cell subsets prior vaccination were positively correlated with the antibody levels in all patients (Roche: ρ = 0.739, R^2^ = 0.001, P<0.001; Table 3; Figure 2B). In this analysis, the absolute number of naïve B cells showed the highest correlation with antibody titers (Roche: ρ = 0.761, R^2^ = 0.153; Figure 2C). Furthermore, this analysis also indicated that the time between last B-cell-depleting therapy and vaccination was also a significant factor correlating with antibody levels. Extending this correlation analysis to the entire study population (Supplementary Figure 2), the influence of naïve B cells remained highly significant (Roche: ρ = 0.636, P<0.001, R^2^ = 0.123).

**Table 3.** Correlations of SARS-CoV-2 antibody response with variables in the immunocompromised study population. Correlations were computed with Spearman’s rank-based rho adjusted for multiple testing (n=21) with Šidák correction. The Šidák-adjusted α level is approximately 0.00244. Significant P-values are highlighted in bold type. Abs., absolute count.

| Variable | Roche anti-SARS-CoV-2 |  |  |
| --- | --- | --- | --- |
| | $\rho$ | P | R<sup>2</sup> |
| DiaSorin SARS-CoV-2 TrimericS IgG | 0.915 | **<0.001** | 0.841 |
| Age | 0.091 | 0.321 | 0.001 |
| Body mass index | 0.169 | 0.076 | 0.004 |
| Days since last B-cell depletion | 0.595 | **<0.001** | 0.041 |
| Interval in days from last B-cell depleting therapy to vaccination up to 365 days | 0.481 | **0.001** | 0.096 |
| IgA | 0.042 | 0.651 | 0.015 |
| IgG | 0.065 | 0.481 | 0.031 |
| IgM | 0.386 | **<0.001** | 0.002 |
| Lymphocytes abs. | 0.222 | 0.018 | 0.002 |
| CD3<sup>+</sup> cells abs. | 0.112 | 0.236 | 0.034 |
| CD3<sup>+</sup>CD8<sup>+</sup> cells abs. | 0.170 | 0.071 | 0.005 |
| CD3<sup>+</sup>CD4<sup>+</sup> cells abs. | 0.025 | 0.789 | 0.062 |
| CD3<sup>-</sup>CD16<sup>+</sup>CD56<sup>+</sup> NK cells abs. | -0.014 | 0.880 | 0.014 |
| CD19<sup>+</sup> abs. | 0.739 | **<0.001** | 0.001 |
| CD45<sup>+</sup> cells abs. | 0.227 | 0.015 | 0.002 |
| CD19<sup>+</sup>IgM<sup>+</sup>CD38<sup>++</sup> transitional B cells abs. | 0.491 | **<0.001** | 0.033 |
| CD19<sup>+</sup>IgD<sup>+</sup>CD27<sup>-</sup> naïve B cells abs. | 0.761 | **<0.001** | 0.153 |
| CD19<sup>+</sup>IgD<sup>+</sup>CD27<sup>+</sup> pre-switched memory B cells abs. | 0.657 | **<0.001** | 0.004 |
| CD19<sup>+</sup>IgD<sup>-</sup>CD27<sup>+</sup> switched memory B cells abs. | 0.710 | **<0.001** | 0.003 |
| CD19<sup>+</sup>CD38<sup>-</sup>CD21<sup>-</sup> B cells abs. | 0.640 | **<0.001** | 0.001 |
| CD19<sup>+</sup>IgM<sup>-</sup>CD38<sup>++</sup> plasmablasts abs. | 0.580 | **<0.001** | 0.001 |

To support the results of our correlation analyses, we established a model of stringent vaccination response prediction for the entire study population based on the results of the Roche assay using the area under the ROC curve (AUC; Supplementary Table 2). This model suggests that the total number of CD19^+^ B cells (AUC: 0.87, 95% CI: 0.79–0.92), CD19^+^IgD^+^CD27^-^ naïve B cells (AUC: 0.82, 95% CI: 0.73–0.88), CD19^+^IgD^+^CD27^+^ pre-switched memory B cells (AUC: 0.84, 95% CI: 0.76–0.89), and CD19^+^IgD^-^CD27^+^ switched memory B cells (AUC: 0.85, 95% CI: 0.79–0.91) can predict response to vaccination in the entire study population.

### CD19^+^IgD^+^CD27^-^ naïve B cells show the strongest association with stringent response

To test the association of variables with vaccination response determined by the Roche assay, we performed univariate and multivariate linear regression analyses using the same variables from the correlation analyses.

In univariate analysis (Table 4), the variables found to be significantly associated with any antibody response in patients included interval in days from the last B-cell-depleting therapy, CD19^+^IgD^+^CD27^-^ naïve B-cell count, CD19^+^IgD^+^CD27^+^ pre-switched memory B-cell count, and absolute number of CD19^+^IgM^-^CD38^++^ plasmablasts. The same correlation was also found for stringent antibody responses, except for the correlation with CD19^+^IgD^+^CD27^+^ pre-switched memory B cells. The absolute number of these B-cell subsets was significantly lower in immunocompromised patients than in healthy controls, in patients after anti-CD20 antibody therapy compared to HSCT, and in patients without seroconversion compared to those with any antibody response (not shown) or stringent response (Supplementary Figure 3). Immunoglobulin levels and cell counts of immunodeficient patients stratified by vaccination response are shown in Supplementary Table 3. In univariate analysis of the entire study population (Supplementary Table 4), the same B-cell subsets remained significantly associated with vaccine response.

**Table 4.**
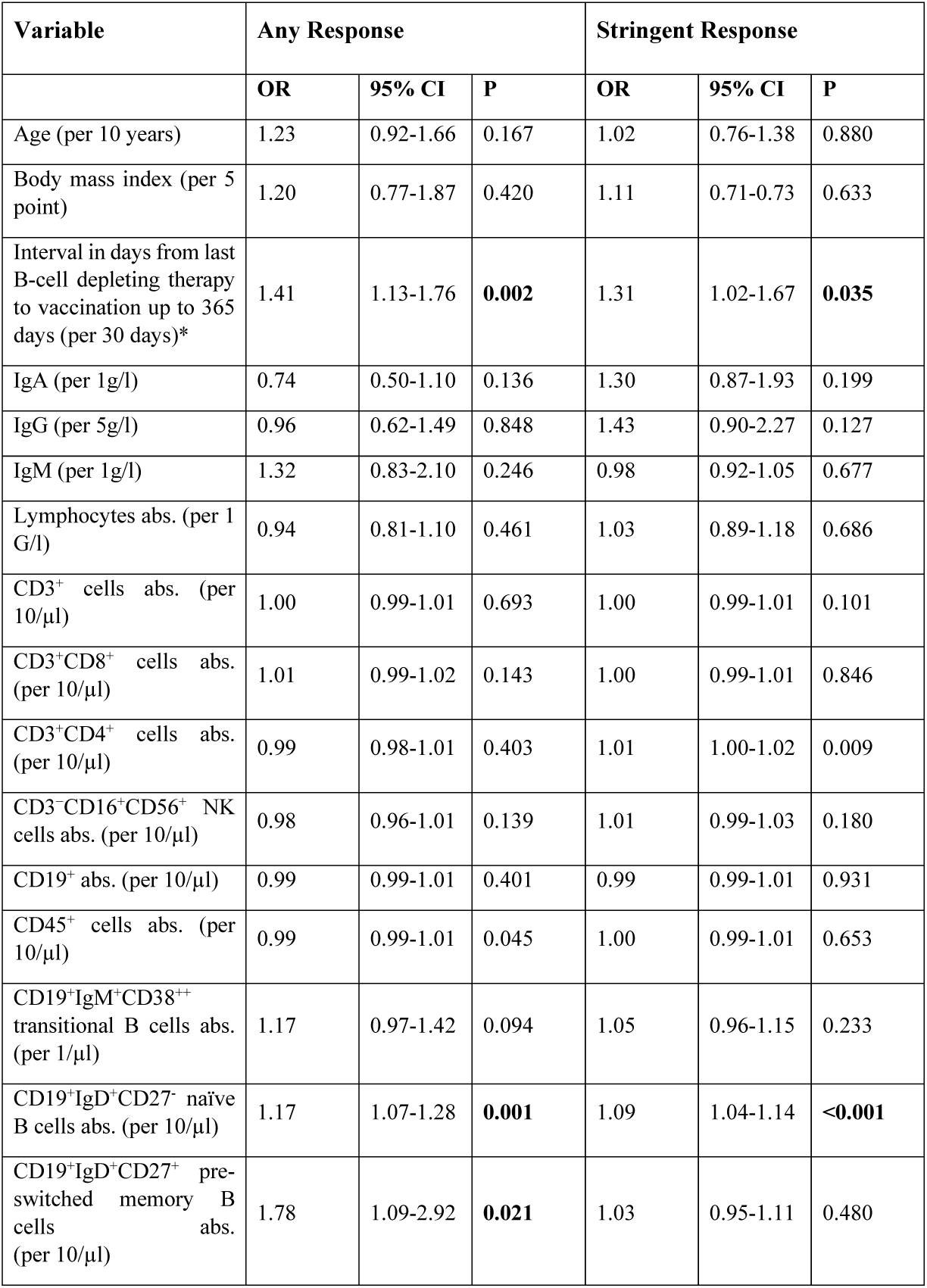
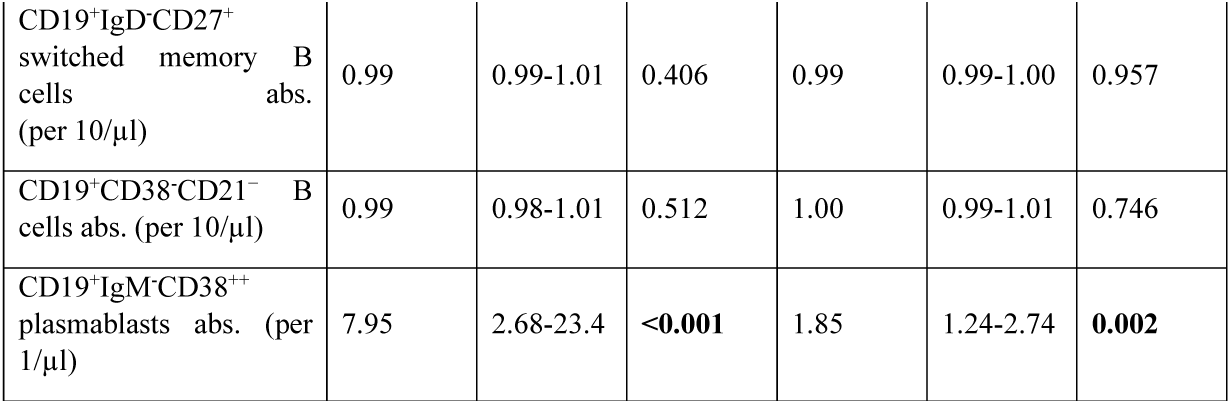
Univariate linear regression analysis to test the association of variables with vaccination response determined by the Roche anti-SARS-CoV-2 S assay in the immunodeficient cohort. Significant P-values are highlighted in bold type. Any response, any seroconversion; OR, odds ratio; stringent response, total antibody titer ≥1000 U/ml. Abs., absolute count.

In multivariable analysis for stringent response, only the number of CD19^+^IgD^+^CD27^-^ naïve B cells was an independent predictor (OR: 1.07 per 10-µl increase, 95% CI: 1.02–1.12, P = 0.009). The only independent predictor of any seroconversion was CD19^+^IgM^-^CD38^++^ plasmablast count (OR: 4.42 per 1-µl increase, 95% CI: 1.30–15.01, P = 0.017).

The multivariable analyses for the entire study population are shown in Supplementary Tables 5 and 6. The absolute number of CD19^+^IgD^+^CD27^-^ naïve B cells remained an independent predictor of stringent antibody response (OR: 1.14 per 10-µl increase, 95% CI: 1.08–1.20, P<0.001).

### Exploratory analyses

As CD19^+^IgD^+^CD27^-^ naïve B cells were the only B-cell subset independently associated with stringent antibody response, we were interested to determine whether our dataset allowed to estimate a CD19^+^IgD^+^CD27^-^ naïve B cell count threshold for stringent antibody response. ROC analysis and non-linear risk modeling predicted that ≥61 CD19^+^IgD^+^CD27^-^ naïve B cells per µl discriminated best between patients with and without a stringent vaccine response (Figures 3A, B).

**Fig 3.**
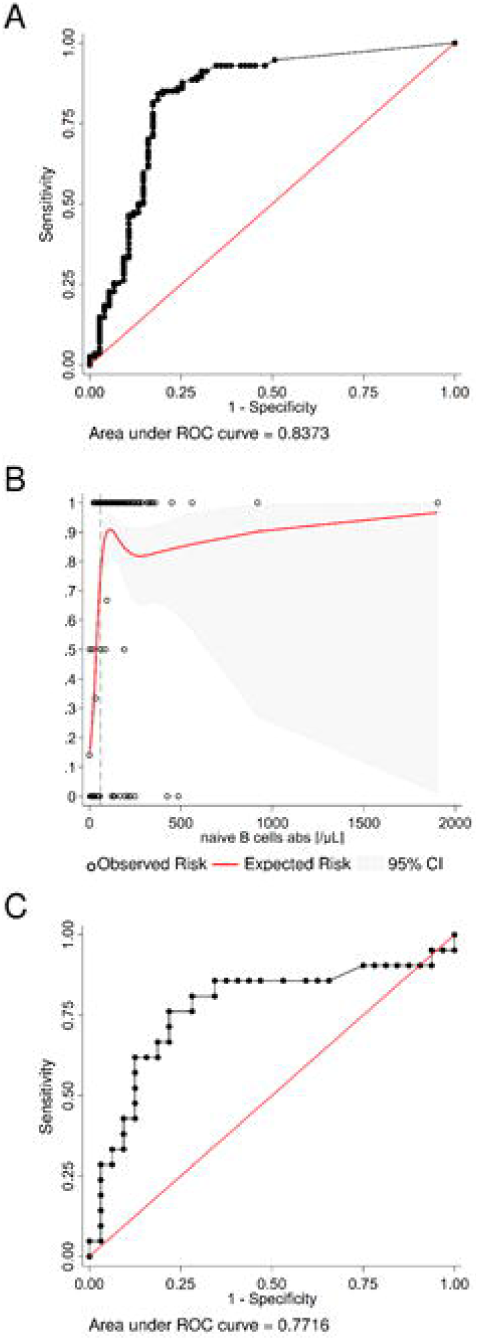
Exploratory analyses estimating the number of naïve B cells and the interval to the last B-cell depleting therapy required for a vaccination response. (A) The receiver operating characteristic (ROC) analysis curve for naïve B cells differentiating stringent antibody response vs no or any vaccination response in the whole study population (n=199) shows excellent discrimination. (B) A non-linear risk model based on the observed risk of seroconversion was created to estimate the minimum number of naïve B cells required for a stringent vaccination response. Independently, the best discriminatory cut-off (dashed line) was determined with the Youden’s index from the ROC curve. Both models predict that ≥60 naïve B cell per µl are required to generate a stringent vaccine response. (C) The ROC analysis curve for the interval since the last B-cell depleting therapy up to 365 days (n=53) differentiating any seroconversion vs no vaccination response. The optimal cut-off is an interval of 116 days or more.

A second exploratory ROC analysis was undertaken to determine the minimum interval between last B-cell-depleting therapy, i.e., anti-CD20 antibody therapy or HSCT up to 365 days (n = 53), and vaccination differentiating any seroconversion versus no vaccination response. The optimal cut-off in this population was 116 days or more (Figure 3C).

## DISCUSSION

Since the presence of B cells is a prerequisite for humoral vaccination responses, we investigated B cells overall and multiple B-cell subsets in immunocompromised patients and healthy controls prior to COVID-19 mRNA vaccination.

Our findings confirm recent reports observing significantly lower and more heterogeneous anti-SARS-CoV-2 S protein IgG titers in immunocompromised patients compared to healthy controls.(14, 15, 22, 24-29) In one of the largest studies, Maneikis and colleagues reported lower median anti-S1 IgG responses after two BNT162b2 vaccine doses in 653 patients with HM compared to 69 healthy healthcare workers.(23) A similar heterogeneity in vaccination response has been observed in patients with PID,(14) AIRD,(16) and individuals given anti-CD20 therapy.(22, 23, 27, 32) In view of the heterogeneity of anti-SARS-CoV-2 antibody levels and the current lack of knowledge regarding the clinical consequences of low versus high titers in immunocompromised patients, we additionally analyzed stringent vaccination response defined as the lowest anti-SARS-CoV-2 antibody titers observed in our healthy control assuming that these would be protective. Based on this definition, only 32% (Roche) and 22% (DiaSorin) of our patients demonstrated a stringent antibody response, respectively. Longitudinal studies are on their way to assess durability of anti-SARS-CoV-2 humoral responses as well as incidences of COVID-19 infections in patients with or without stringent humoral response.

In our study, absolute numbers of CD19^+^IgD^+^CD27^-^ naïve B cells, CD19^+^IgD^+^CD27^+^ pre-switched memory B cells, and CD19^+^IgM^-^CD38^++^ plasmablasts were significantly associated with anti-SARS-CoV-2 response in univariate analysis. However, only the number of CD19^+^IgD^+^CD27^-^ naïve B cells was an independent predictor of a stringent vaccination response in multivariable analysis, suggesting their functional importance for obtaining a humoral immune response. Indeed, the production of specific antibodies to a novel antigen relies on the presence of antigen-specific B cells within the CD19^+^IgD^+^CD27^-^ naïve B cell population.(33) Thus, a drastically contracted pool of CD19^+^IgD^+^CD27^-^ naïve B cells reduces the chance of harboring B cells with a B-cell receptor of high antigen avidity that can interact with T follicular helper cells successfully and subsequently undergo somatic hypermutation to develop an optimal antibody response.(34) A lack of these cells results in low antibody titers of poor quality. Therefore, the association of the magnitude of the humoral vaccination response with the abundance of CD19^+^IgD^+^CD27^-^ naïve B cells is most likely a causal relationship.

To date, there is little literature on this topic, and we are the first to describe this relationship, particularly in the context of COVID-19 and immunocompromised patients. The importance of naïve B cells for antibody response has already been shown for the H5N1 influenza vaccine, but to the best of our knowledge not yet for immunocompromised patient cohorts or COVID-19 in particular.(35) Recently, Redjoul and colleagues reported a significant increase in anti-SARS-CoV-2 antibodies in 42 HM patients given a third dose of BNT162b2 vaccine after allogeneic HSCT.(36) In a multivariable analysis, only a peripheral B-cell count of more than 250/µl at the time of the third vaccination was associated with humoral response (OR 7.1, 95% CI: 1.5-34.1, P=0.016).(36) Mrak et al. observed that the percentage of peripheral CD19^+^ B cells positively correlated with antibody levels after BNT162b2 vaccination (τ=0.4, P<0.001) in patients with AIRD after rituximab therapy.[24] The median percentage of peripheral CD19^+^ B cells was 2% (interquartile range [IQR], 0-33) in the study of Mrak et al., which is not very different from 4% (IQR, 0-13) in our patient population. Our results extend these findings and show that only CD19^+^IgD^+^CD27^-^ naïve B cells predicted a strong vaccination response.

How soon humoral immune responses may be expected after B-cell depleting therapy remains a concern, and recommendations of medical societies differ. Our data confirm that the interval between the last B-cell-depleting therapy and SARS-CoV-2 vaccination plays a crucial role in achieving seroconversion for immunocompromised patients. We were able to define a minimum of 116 days, i.e. 4 months, from last B-cell-depleting therapy to vaccination as prerequisite for obtaining an anti-SARS-CoV-2 seroconversion.

Whereas patients after allogeneic HSCT have high numbers of CD19^+^IgD^+^CD27^-^ naïve B cells, this B-cell subpopulation was significantly lower in individuals suffering from chronic graft-versus-host disease (GVHD).(37) Long-term clinical efficacy of rituximab could be demonstrated in chronic GVHD patients recovering naïve B cells after treatment. These findings are consistent with clinical responses to rituximab reported in patients with rheumatoid arthritis, SLE, and mixed cryoglobulinemia who recovered B cells.(38-40) Thus, rise in absolute numbers of B cells as well as CD19^+^IgD^+^CD27^-^ naïve B cells might indicate immune reconstitution after immunosuppressive therapies enabling achievement of a humoral immune response to SARS-CoV-2 vaccination. Contrarily, patients with a deficit of CD19^+^IgD^+^CD27^-^ naïve B cells may benefit more from continuation of strict hygiene measures as recommended by scientific organizations. Whether these immunocompromised patients will substantially benefit from a third vaccination, must be demonstrated since first data of HSCT patients showed only low antibody titers in half of the patients.(36)

Our study had several limitations. These include single-center design and limited representation of some patient cohorts that do not allow clear conclusions on seroconversion rates among less common entities or less frequently used treatment strategies. Moreover, we cannot comment on the persistence of the observed vaccination response at this point. Our study relies on the measurement of antibodies as a surrogate for immunity to SARS-CoV-2. However, our results do not significantly differ between the two internationally deployed anti-spike protein serological assays for detecting either total Ig or IgG. Both tests showed a high correlation with surrogate neutralization tests, and Roche’s assay correlated well with live virus neutralization tests in vaccinated individuals.(29, 31) The strength of our study is its prospective design and the introduction of the stringent vaccination response as a potentially clinically more relevant concept than seroconversion.

In summary, humoral response to SARS-CoV-2 mRNA vaccine is impaired in immunocompromised patients. The abundance of circulating CD19^+^IgD^+^CD27^-^ naïve B cells is strongly associated with an improved antibody vaccine response across different diseases and therapies. Therefore, measuring CD19^+^IgD^+^CD27^-^ naïve B cells may allow prediction of a humoral response to COVID-19 vaccination in immunocompromised patients. Further research is needed to confirm these findings for vaccinating immunocompromised individuals against COVID-19 and other pathogens.

## Data Availability

All data will be shared upon request.

## Acknowledgments

We thank Pia-Carina Gallaun and Julia Lodron for technical assistance. The samples/data used for this project were processed with the help of the Biobank Graz of the Medical University of Graz, Austria.

## Author contributions

IH, ES, PS, SH, HG, IS, and MHS designed the study. ES, IH, PF, NS, JF, BD, BU, AG, MM, CU, MK, CE and BK collected clinical samples and/or data. PF and BK performed antibody assays. ES and SH analyzed the data and performed the statistical analysis. ES, SH, IH, PS, HG, IS, and MHS critically reviewed and discussed the results. ES, IH, PF, HS, HG, and MHS wrote the first draft. All authors reviewed the draft and approved the final version of the manuscript.

