## Supplemental tables and figures for "CD19^+^IgD^+^CD27^-^ naïve B Cells as predictors of humoral response to COVID-19 mRNA vaccination in immunocompromised patients"

### Table Legends.

**Supplementary Table 1. Diagnoses and immunosuppressive agents stratified by vaccination response determined by the Roche anti-SARS-CoV-2 S assay.** Any response, any seroconversion; GVHD, graft-versus-host-disease; HSCT, hematopoietic stem cell transplantation; IMiD, immunomodulatory imide drug; stringent response, total antibody titre ≥1000 U/mL.

**Supplementary Table 2. Model of COVID-19 vaccination response prediction for the whole study population.** Response was measured with the Roche anti-SARS-CoV-2 S assay. The predictive ability of different variables is indicated by the area under the receiver operating characteristic curve. The bootstrap method was used to determine 95% confidence intervals. AUC ≥ 0.8 were considered to show excellent discrimination and highlighted in bold type. Abs., absolute; any response, any seroconversion; AUC, area under curve; stringent response, total antibody titer ≥1000 U/mL.

**Supplementary Table 3. Variables stratified by vaccination response determined by the Roche anti-SARS-CoV-2 S assay in the patient population.** Data are reported as medians [25th-75th percentile]. Any response, any seroconversion; stringent response, total antibody titer ≥1000 U/mL.

**Supplementary Table 4. Univariate linear regression analysis to test the association of variables with vaccination response determined by the Roche anti-SARS-CoV-2 S assay in the complete study population.** Significant P-values are highlighted in bold type. Abs., absolute; any response, any seroconversion; OR, odds ratio; stringent response, total antibody titer ≥1000 U/mL.

**Supplementary Table 5. Multiple linear regression analysis to test the independent effect of variables on seroconversion determined by the Roche anti-SARS-CoV-2 S assay in the complete study population.** Variables were selected from the univariate linear regression analysis and tested stepwise. Interval to B-cell depleting therapy was excluded because this variable was not applicable to all patients. Significant P-values are highlighted in bold type. Abs., absolute; any response, any seroconversion; OR, odds ratio.

**Supplementary Table 6. Multiple linear regression analysis to test the independent effect of variables on stringent vaccination response as determined by the Roche Anti-SARS-CoV-2 S assay in the complete study population.** Variables were selected from the univariate linear regression analysis. Interval to B-cell depleting therapy was excluded because this variable was not applicable to all patients. Significant P-values are highlighted in bold type. Abs., absolute; OR, odd ratio; stringent response, total antibody titer ≥1000 U/mL.

### Supplementary Table 1.

| | Overall n | No response n | Any response n | Stringent response n | P |
|---|---|---|---|---|---|
| **Diagnostic group** | **120** | **40 (33%)** | **80 (67%)** | **38 (32%)** | **<0.001** |
| **Primary immunodeficiency** | 25 | 2 (8%) | 23 (92%) | 16 (64%) | |
| Antibody and/or subclass deficiency | 12 | 0 | 12 (10%) | 11 (92%) | |
| Common variable immunodeficiency | 11 | 2 (18%) | 9 (82%) | 4 (36%) | |
| Other | 2 | 0 | 2 (100%) | 1 (50%) | |
| **Autoimmune diseases** | 39 | 30 (77%) | 9 (23%) | 3 (8%) | |
| Connective tissue disease | 22 | 18 (82%) | 4 (18%) | 1 (5%) | |
| Multiple sclerosis spectrum disorder | 8 | 6 (75%) | 2 (25%) | 0 | |
| Other | 4 | 3 (75%) | 1 (25%) | 0 | |
| Rheumatoid arthritis | 5 | 3 (60%) | 2 (40%) | 2 (40%) | |
| **Hematologic diseases*** | 56 | 8 (14%) | 48 (86%) | 19 (34%) | |
| Acute myeloid leukemia | 14 | 2 (14%) | 12 (86%) | 7 (50%) | |
| GVHD after allogeneic HSCT | 14 | 2 (14%) | 12 (86%) | 7 (50%) | |
| Lymphoid neoplasms | 13 | 5 (38%) | 8 (62%) | 4 (31%) | |
| Multiple myeloma[#] | 15 | 0 | 15 (100%) | 4 (27%) | |
| Myelodysplastic syndrome | 5 | 1 (20%) | 4 (80%) | 0 | |
| Myeloproliferative neoplasms | 4 | 0 | 4 (100%) | 2 (50%) | |
| Other | 6 | 0 | 6 (100%) | 2 (33%) | |
| **Immunosuppressive medication** | | | | | |
| -Any | 89 | 37 (42%) | 52 (58%) | 18 (20%) | **<0.001** |
| -None | 31 | 3 (10%) | 28 (90%) | 20 (65%) | |
| Abatacept | 1 | 0 | 1 (100%) | 0 | |
| Anti-CD20 | 41 | 31 (76%) | 10 (24%) | 4 (10%) | |
| -Ocrelizumab | 6 | 5 (83%) | 1 (17%) | 0 | 0.631 |

| | | | | | |
|---|---|---|---|---|---|
| -Rituximab | 35 | 26 (74%) | 9 (27%) | 4 (10%) | |
| Azacitidine | 2 | 0 | 2 (100%) | 0 | |
| Azathioprine | 1 | 0 | 1 (100%) | 1 (100%) | |
| Bendamustine | 2 | 1 (50%) | 1 (50%) | 0 | |
| Ciclosporin | 15 | 2 (13%) | 13 (87%) | 5 (33%) | |
| Daratumumab | 9 | 0 | 9 (100%) | 1 (11%) | |
| Ibrutinib | 1 | 0 | 1 (100%) | 0 | |
| IMiD | 11 | 0 | 11 (100%) | 2 (18%) | |
| Mycophenolate mofetil/ mycophenolic acid | 9 | 8 (89%) | 1 (11%) | 0 | |
| Proteasome inhibitor | 6 | 0 | 6 (100%) | 3 (50%) | |
| Ruxolitinib | 6 | 2 (33%) | 4 (67%) | 2 (33%) | |
| Steroid monotherapy | 2 | 0 | 2 (100%) | 1 (50%) | |
| Venetoclax | 1 | 1 (100%) | 0 | 0 | |
| Hematopoietic stem cell transplantation | 35 | 4 (11%) | 31 (89%) | 13 (37%) | |
| -Allogeneic | 25 | 4 (16%) | 21 (84%) | 10 (40%) | 0.366 |
| -Autologous | 10 | 0 | 10 (100%) | 3 (30%) | |

*Some patients have multiple diagnoses; #all under active treatment.

### Supplementary Table 2.

| Variable | Any Response | | Stringent Response | |
|---|---|---|---|---|
| | **AUC** | **95% CI** | **AUC** | **95% CI** |
| Age | 0.46 | 0.36-0.58 | 0.36 | 0.28-0.45 |
| Body mass index | 0.53 | 0.43-0.64 | 0.46 | 0.37-0.54 |
| IgA | 0.54 | 0.43-0.46 | 0.68 | 0.59-0.76 |
| IgG | 0.58 | 0.46-0.69 | 0.66 | 0.57-0.74 |
| IgM | 0.68 | 0.58-0.77 | 0.76 | 0.68-0.83 |
| Lymphocytes abs. | 0.46 | 0.27-0.64 | 0.65 | 0.57-0.72 |
| $CD3^{+}$ cells abs. | 0.58 | 0.48-0.68 | 0.61 | 0.52-0.70 |
| $CD3^{+}CD8^{+}$ cells abs. | 0.61 | 0.51-0.72 | 0.54 | 0.44-0.63 |
| $CD3^{+}CD4^{+}$ cells abs. | 0.51 | 0.41-0.61 | 0.67 | 0.59-0.75 |
| $CD3^{-}CD16^{+}CD56^{+}$ NK cells abs. | 0.45 | 0.35-0.57 | 0.54 | 0.45-0.63 |
| $CD19^{+}$ abs. | **0.92** | 0.82-0.98 | **0.87** | 0.79-0.92 |
| $CD45^{+}$ abs. | 0.64 | 0.55-0.74 | 0.68 | 0.60-0.76 |
| Transitional B cells abs. | 0.65 | 0.56-0.73 | 0.60 | 0.50-0.69 |
| Naïve B- cells abs. | **0.88** | 0.79-0.94 | **0.82** | 0.73-0.88 |
| $IgD^{+}$ memory B cells abs. | **0.84** | 0.76-0.91 | **0.83** | 0.76-0.89 |
| $IgD^{-}$ memory B cells abs. | **0.87** | 0.81-0.93 | **0.85** | 0.79-0.91 |
| $CD21^{-}$ B cells abs. | **0.86** | 0.77-0.93 | 0.76 | 0.67-0.84 |
| Plasmablasts abs. | 0.73 | 0.64-0.81 | 0.69 | 0.59-0.77 |

### Supplementary Table 3.

| Variable | Overall (n=120) | No response (n=40) | Any response (n=42) | Stringent response (n=38) |
|---|---|---|---|---|
| Age (years) | 58.0 [50.0-65.1] | 54.4 [43.1-65.2] | 61.1 [54.3-65.3] | 58.0 [50.9-65.1] |
| Body mass index (kg/m²) | 24.8 [22.7-27.8] | 24.1 [22.1-26.8] | 25.0 [23.7-28.3] | 25.4 [23.2-28.9] |
| Days since anti-CD20 therapy* | 69 [46-132] | 62.0 [41-100] | 129 [51-163] | 222 [157 -288] |
| IgA (g/L) | 1.1 [0.4-1.8] | 1.3 [0.5-2.1] | 0.5 [0.1-1.4] | 1.37 [0.7-1.8] |
| IgG (g/L) | 8.2 [6.2-10.6] | 8.7 [6.3-11.1] | 7.6 [5.7-.9.4] | 8.9 [7.0-11.2] |
| IgM (g/L) | 0.4 [0.2-0.8] | 0.4 [0.2-0.7] | 0.3 [0.2-0.6] | 0.6 [0.4-1.1] |
| Lymphocytes (x1000/µL) | 1.4 [0.9-1.9] | 1.3 [0.9-1.6] | 1.3 [0.8-2.4] | 1.8 [1.1-2.4] |
| $CD3^+$ cells (x1000/µL) | 1.1 [0.7-1.5] | 1.0 [0.7-1.2] | 0.9 [0.5-1.5] | 1.2 [0.7-1.6] |
| $CD3^+CD8^+$ cells (x1000/µL) | 0.4 [0.3-0.7] | 0.3 [0.2-0.6] | 0.4 [0.2-0.9] | 0.5 [0.3-0.7] |
| $CD3^+CD4^+$ cells (x1000/µL) | 0.5 [0.3-0.8] | 0.6 [0.4-0.8] | 0.4 [0.2-0.6] | 0.7 [0.4-1.1] |
| $CD3^-CD16^+56^+$ NK cells (x1000/µL) | 0.2 [0.1-0.3] | 0.2 [0.2-0.3] | 0.2 [0.1-0.3] | 0.2 [0.2-0.3] |
| $CD19^+$ cells (/µL) | 47 [0-170] | 0 [0-0] | 58 [16-128] | 186 [115-332] |
| $CD45^+$ cells (x1000/µL) | 1.4 [0.9-1.9] | 1.3 [0.9-1.5] | 1.3 [0.8-1.7] | 1.7 [1.2-2.4] |
| Transitional B cells (/µL) | 0 [0-2] | 0 [0-0] | 0 [0-2] | 1 [1-3] |
| Naïve B cells (/µL) | 18 [0-133] | 0 [0-0] | 34 [7-96] | 144 [74-221] |
| $IgD^+$ memory B cells (/µL) | 2 [0-10] | 0 [0-0] | 3 [1-9] | 9 [3-34] |
| $IgD^-$ memory B cells (/µL) | 2 [0-15] | 0 [0-0] | 4 [2-12] | 18 [4-30] |
| $CD21^-$ B cells (/µL) | 5 [0-19] | 0 [0-0] | 6 [2-22] | 17 [5-36] |
| Plasmablasts (/µL) | 0 [0-1] | 0 [0-0] | 0 [0-1] | 1 [1-2] |

*Anti-CD20 therapy within 365.25 days.

### Supplementary Table 4.

| Variable | Any response | | | Stringent response | | |
|---|---|---|---|---|---|---|
| | OR | 95% CI | P | OR | 95% CI | P |
| Age (per 10 years) | 0.90 | 0.69-1.17 | 0.429 | 0.69 | 0.54-0.87 | **0.001** |
| Body mass index (per 5 point) | 1.04 | 0.86-1.27 | 0.688 | 1.03 | 0.94-1.13 | 0.565 |
| Interval in days from last B-cell depleting therapy to vaccination up to 365 days (per 30 days)* | 1.41 | 1.13-1.76 | **0.002** | 1.31 | 1.02-1.67 | **0.035** |
| IgA (per 1g/L) | 1.15 | 0.80-1.67 | 0.451 | 2.15 | 1.51-3.05 | **<0.001** |
| IgG (per 5g/L) | 1.52 | 0.89-2.58 | 0.121 | 3.00 | 1.79-5.04 | **<0.001** |
| IgM (per 1g/L) | 2.45 | 1.17-5.13 | **0.017** | 0.98 | 0.93-1.04 | 0.512 |
| Lymphocytes abs. (per 1 G/L) | 0.93 | 0.81-1.08 | 0.343 | 1.01 | 0.87-1.17 | 0.867 |
| $CD3^+$ cells abs. (per 10/µL) | 1.00 | 0.99-1.01 | 0.512 | 1.00 | 0.99-1.01 | 0.071 |
| $CD3^+CD8^+$ cells abs. (per 10/µL) | 1.00 | 0.99-1.01 | 0.309 | 0.99 | 0.98-1.00 | 0.417 |
| $CD3^+CD4^+$ abs. (per 10/µL) | 0.99 | 0.99-1.01 | 0.975 | 1.02 | 1.00-1.03 | **<0.001** |
| $CD3^-CD16^+56^+$ NK cells abs. (per 10/µL) | 0.98 | 0.96-1.01 | 0.117 | 1.01 | 0.99-1.03 | 0.232 |
| $CD19^+$ cells abs. (per 10/µL) | 0.99 | 0.99-1.01 | 0.272 | 0.99 | 0.99-1.00 | 0.651 |
| $CD45^+$ cells abs. (per 10/µL) | 0.99 | 0.99-1.01 | 0.337 | 1.00 | 0.99-1.00 | 0.823 |
| Transitional B cells abs. (per 1/µL) | 1.29 | 0.98-1.70 | 0.066 | 1.03 | 0.95-1.12 | 0.416 |
| Naïve B cells abs. (per 10/µL) | 1.26 | 1.14-1.39 | **<0.001** | 1.15 | 1.09-1.20 | **<0.001** |
| $IgD^+$ memory B cells abs. (per 10/µL) | 3.24 | 1.83-5.73 | **<0.001** | 1.30 | 1.09-1.55 | **0.004** |
| $IgD^-$ memory B cells abs. (per 10/µL) | 0.99 | 0.99-1.01 | 0.321 | 0.99 | 0.99-1.00 | 0.425 |
| $CD21^-$ B cells abs. (per 10/µL) | 0.99 | 0.98-1.01 | 0.270 | 0.99 | 0.99-1.01 | 0.686 |
| Plasmablasts abs. (per 1/µL) | 13.33 | 4.66-38.01 | **<0.001** | 2.85 | 1.88-4.32 | **<0.001** |

* Includes all patients after anti-CD20 therapy or HSCT (n=53).

### Supplementary Table 5.

| Multivariable Model | Variable | Multivariable OR | 95%CI | P |
|---|---|---|---|---|
| #1 (n=189) | Naïve B cells abs. (per 10/µL) | 1.25 | 1.13-1.38 | **<0.001** |
| | IgM (per 1g/L) | 0.33 | 0.75-2.37 | 0.336 |
| #2 (n=189) | Naïve B cells abs. (per 10/µL) | 1.18 | 1.06-1.31 | **0.003** |
| | $IgD^{+}$ memory B cells abs. (per 10/µL) | 1.56 | 0.95-2.56 | 0.078 |
| #3 (n=189) | Naïve B cells abs. (per 10/µL) | 1.13 | 1.01-1.25 | 0.028 |
| | Plasmablasts abs. (per 1/µL) | 5.68 | 1.77-18.18 | **0.003** |
| #4 (n=189) | Naïve B cells abs. (per 10/µL) | 1.10 | 0.98-1.23 | 0.094 |
| | IgM (per 1g/L) | 1.13 | 0.67-1.92 | 0.641 |
| | $IgD^{+}$ memory B cells abs. (per 10/µL) | 1.22 | 0.78-1.92 | 0.376 |
| | Plasmablasts abs. (per 1/µL) | 4.82 | 1.49-15.63 | **0.009** |

### Supplementary Table 6.

| Multivariable Model | Variable | Multivariable OR | 95%CI | P |
|---|---|---|---|---|
| #1 (n=189) | Naïve B cells abs. (per 10/µL) | 1.14 | 1.08-1.20 | **<0.001** |
| | Age (per 10 years) | 0.85 | 0.65-1.11 | 0.234 |
| #2 (n=189) | Naïve B cells abs. (per 10/µL) | 1.15 | 1.09-1.21 | **<0.001** |
| | IgA (per 1g/L) | 2.22 | 1.48-3.35 | **<0.001** |
| #3 (n=189) | Naïve B cells abs. (per 10/µL) | 1.14 | 1.08-1.20 | **<0.001** |
| | IgG (per 5g/L) | 5.59 | 1.78-17.50 | 0.003 |
| #4 (n=189) | Naïve B cells abs. (per 10/µL) | 1.14 | 1.09-1.20 | **<0.001** |
| | $CD3^+CD4^+$ cells abs. (per 10/µL) | 1.02 | 1.01-1.03 | 0.003 |
| #5 (n=189) | Naïve B cells abs. (per 10/µL) | 1.14 | 1.08-1.20 | **<0.001** |
| | $IgD^+$ memory B cells abs. (per 10/µL) | 1.03 | 0.88-1.20 | 0.732 |
| #6 (n=189) | Naïve B cells abs. (per 10/µL) | 1.15 | 1.09-1.20 | **<0.001** |
| | Plasmablasts abs. (per 1/µL) | 1.00 | 0.90-1.11 | 0.979 |
| #7 (n=189) | Naïve B cells abs. (per 10/µL) | 1.14 | 1.08-1.20 | **<0.001** |
| | Age (per 10 years) | 0.94 | 0.70-1.28 | 0.723 |
| | IgA (per 1g/L) | 1.81 | 1.13-2.88 | **0.013** |
| | IgG (per 5g/L) | 2.76 | 0.74-10.20 | 0.129 |
| | $CD3^+CD4^+$ cells abs. (per 10/µL) | 1.02 | 1.00-1.03 | **0.006** |
| | $IgD^+$ memory B cells abs. (per 10/µL) | 1.00 | 0.89-1.11 | 0.981 |
| | Plasmablasts abs. (per 1/µL) | 1.02 | 0.91-1.14 | 0.764 |

### Figure Legends.

**Supplementary Figure 1**. **Flow cytometry gating strategy for B cell subsets.** A) patient with normal B-cell count: gating of $CD21^{-}$ B cells (second row, left), switched and pre-switch memory B cells (second row, middle), transitional B cells and plasmablasts (second row, right); B) patient with low B-cell count and no further subphenotyping. Arrows indicate consecutive subphenotyping of the cells selected by gates in prior window. SSC-A, side scatter (area); FSC-A, forward scatter (area).

**Supplementary Figure 2**. **Correlation of antibody levels determined by Roche anti-SARS-CoV-2 S assay with variables from univariate linear regression analysis in the complete study population.** (A.) DiaSorin SARS-CoV-2 TrimericS IgG. (B.) Age. (C.) IgA. (D.) IgG. (E.) $CD3^{+}CD4^{+}$ cells. (F.) Naïve B cells. Scatter plots indicate a linear regression line including a 95% confidence interval.

**Supplementary Figure 3**. **Absolute counts of B-cell-subsets of healthy participants and immunodeficient patients as well as according to COVID-19 vaccination response.** (A.) $CD19^{+}$ cells. (B.) Naïve B cells. (C.) $IgD^{+}$ memory B cells. (D.) Plasmablasts. P is <0.05 for all comparisons. Data points were transformed with inverse hyperbolic sine transformation of x to retain zero-valued measurements. White circles are medians, box limits indicate the 25th and 75th percentiles, whiskers extend 1.5 times the interquartile range from the 25th and 75th percentiles, polygons represent density estimates of data and extend to extreme values. Violin plots were drawn with BoxPlotR. Healthy probands, n = 79; immunodeficient patients, n= 110; patients after anti-CD20 therapy, n= 41; patients after HSCT, n=28; immunodeficient patients with no seroconversion, n=38; immunodeficient patients with stringent response, n= 35. IHS, inverse hyperbolic sine transformed; HSCT, hematopoietic stem cell transplantation.

### Supplementary Figure 1.

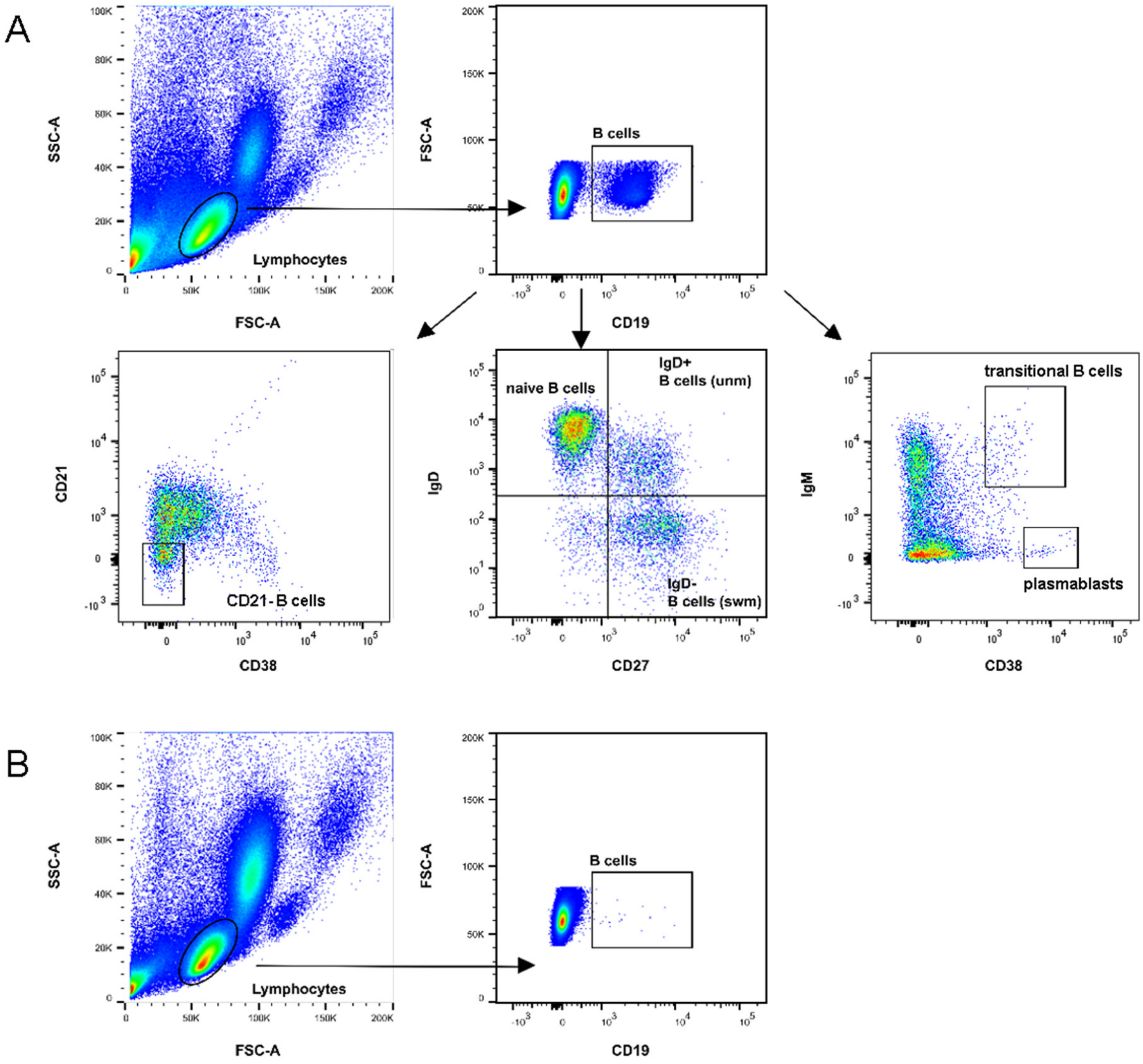

### Supplementary Figure 2.

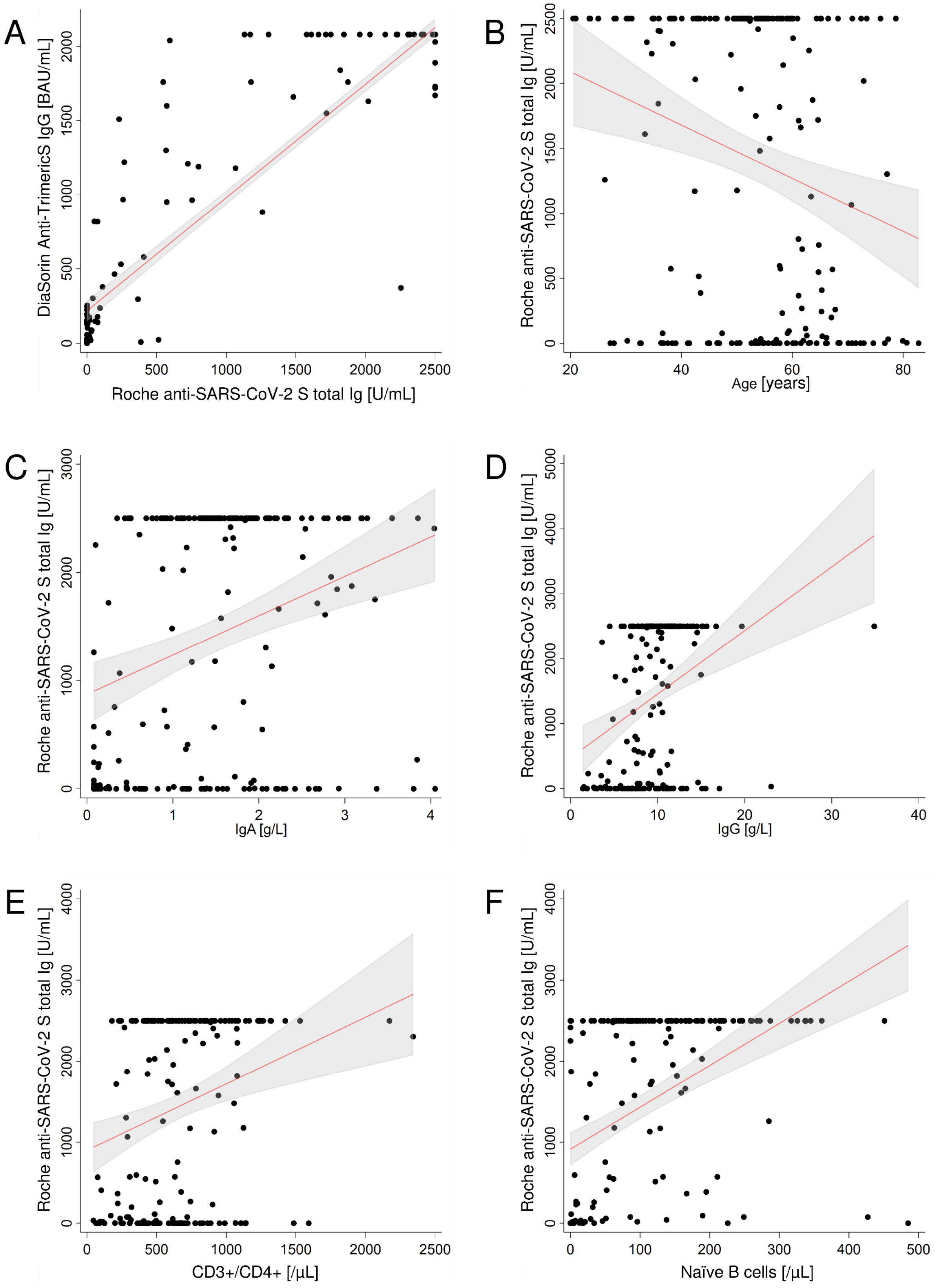

### Supplementary Figure 3.

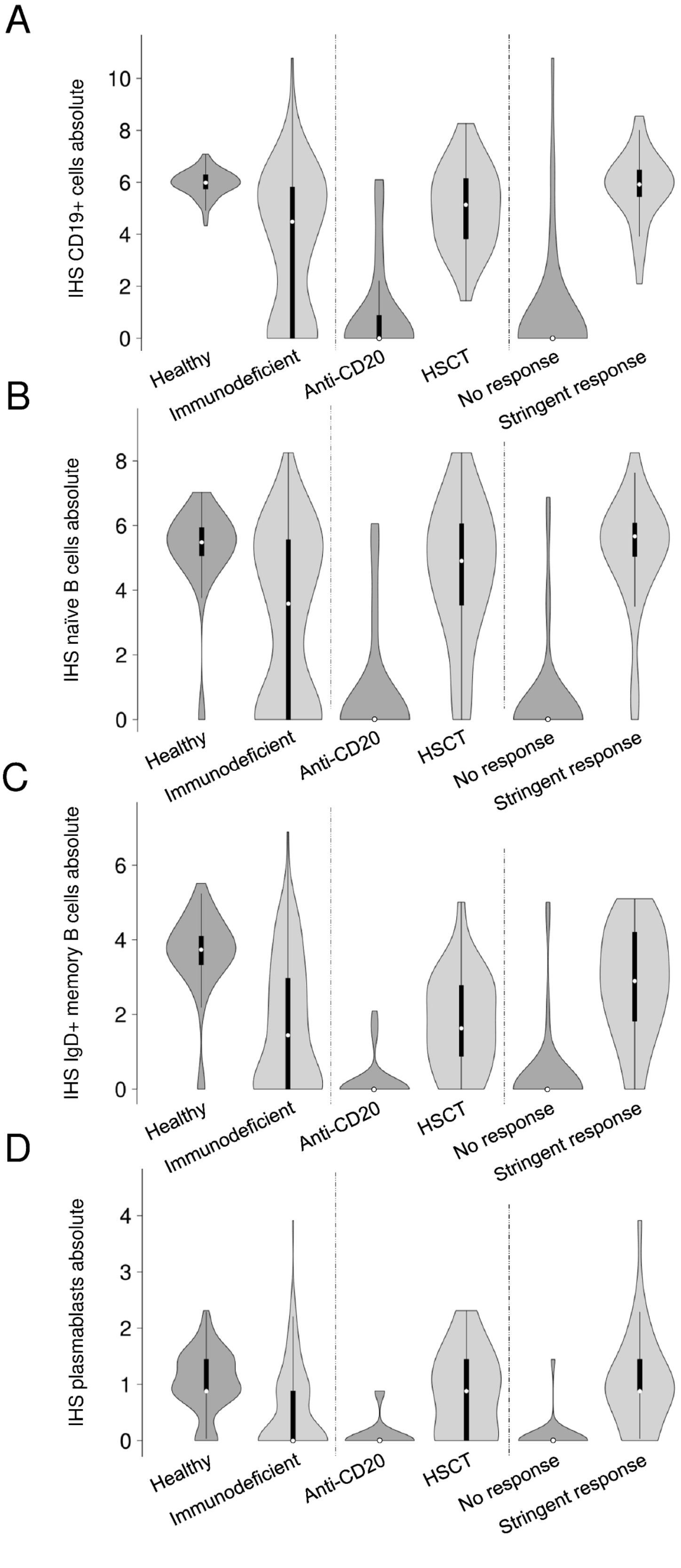

### Humoral and cellular immune response to COVID-19 vaccines in immunocompromised and healthy individuals – The CoVVac study

A prospective monocentric cohort study

Version 3.0, 08.04.2021

Authors:

**Isabel Hodl, Eduard Schulz, Peter Schlenke, Stefan Hatzl, Martin Stradner, Hildegard Greinix, Ivo Steinmetz**

Sponsor: **Medical University of Graz**

| | |
|---|---|
| Sponsor's study protocol code | CoVVac |
| EudraCT number | 2021-001040-10 |

Principal Investigator:

Assoz.Prof. PD Dr. med. univ. Martin Stradner

Medical University Graz

Division of Rheumatology and Immunology

Auenbruggerplatz 15

8036 Graz – Austria

+43 316-385-81794

The information contained in this study protocol is absolutely confidential. Its sole purpose is to inform the sponsor, the investigators, the study staff, the ethics committee, the official authorities, and patients. This study protocol may not be passed on to a third party without the consent of the sponsor or the Principal Investigator of the clinical trial.

#### Contents

Page

**List of abbreviations** ... **4**

**Responsibilities and addresses** ... **7**

**Synopsis** ... **9**

**1. Scientific background** ... **14**

**2. Aim of the trial** ... **21**

2.1. Study design ... 22

2.1.1 Time schedule ... 22

2.2 Outcome measures ... 23

2.2.1 Primary outcome measures ... 23

2.2.2 Secondary outcome measures ... 23

**3. Recruitment of probands** ... **23**

3.1 Number of probands and the duration of the trial ... 23

3.2 Inclusion criteria ... 24

3.3 Exclusion criteria ... 25

3.4 Clinical history ... 26

3.5 Parameters, Laboratory parameters ... 26

**4. Implementation of the trial / Course of the trial** ... **27**

4.1 Description of the individual visits ... 28

4.2 Methods ... 29

4.3 Risk-benefit assessment and precautionary measures ... 34

4.3.1 Pregnancy tests ... 34

4.4 Adverse events ... 34

4.4.1 Definitions ... 34

4.4.2 Reporting obligations ... 35

4.5 Interim analyses ... 35

4.6 Termination of the trial ... 36

4.6.1 Termination of the trial for a subject (drop-out) ... 36

4.6.2 Termination of the entire trial ... 36

**5. Monitoring and Audit – Assurance of data quality** ... **36**

5.1 Monitoring ... 37

5.2 Audit ... 38

**6. Biometry** ... **38**

#### List of abbreviations

| | |
|---|---|
| ACE2 | Angiotensin converting enzyme 2 |
| AE | Adverse event |
| AMG | Arzneimittelgesetz (Austrian Act on Pharmaceutical Products) |
| AR | Adverse reaction |
| BASG | Bundesamt für Sicherheit im Gesundheitswesen (Federal Agency for Safety in Healthcare) |
| BCIP | 5-bromo-4-chloro-3'-indolyl phosphate |
| BMI | Body mass index |
| cDNA | Complementary deoxyribonucleic acid |
| CLL | Chronic lymphocytic leukemia |
| CoV | Coronavirus |
| COVID-19 | Coronavirus disease 2019 |
| CRF | Case report forms |
| CVID | Common variable immunodeficiency |
| DII | Dietary inflammation inde |
| DMSO | Dimethyl sulfoxide |
| E | Envelope protein |
| ECIL | European Conference on Infections in Leukemia |
| EFSA | European Food Safety Authority |
| ELISpot | Enzyme-linked immunosorbent spot |
| EMA | European Medicines Agency |

| | |
|---|---|
| FACS | Fluorescence-activated cell sorting |
| FPFV | First Patient First Visit |
| GCP | Good clinical practice |
| HSCT | Hematopoietic stem-cell transplantation |
| IB | Investigator brochure |
| IBD | Inflammatory bowel disease |
| ICH | International Council on Harmonisation of Technical Requirements for Registration of Pharmaceuticals for Human Use |
| IFNγ | Interferon γ |
| IgA/IgG/IgM | Immunoglobulin A/G/M |
| IMPD | Investigator Medicinal Product Dossier |
| International Society for the Advancement of Kinanthropometry | ISAK |
| LPLV | Last Patient First Visit |
| M | Membrane protein |
| MACS | Magnetic-activated cell sorting |
| MERS-CoV | Middle East respiratory syndrome-related coronavirus |
| MPN | Myeloproliferative neoplasm |
| mRNA | Messenger RNA |
| N | Nucleocapsid protein |
| NBT | Nitro blue tetrazolium |
| PBMCs | Peripheral blood mononuclear cells |
| PCR | Polymerase chain reaction |
| PHM | Patients with hematological malignancies |

| | |
|---|---|
| **RBD** | **Receptor-binding domain** |
| **RDA** | **Research, Documentation & Analysis** |
| **RNA** | **Ribonucleic acid** |
| **S** | **Spike protein** |
| **SAE** | **Serious adverse event** |
| **SAR** | **Serious adverse reaction** |
| **SARS-CoV (-2)** | **severe acute respiratory syndrome coronavirus (type 2)** |
| **SDV** | **Source data verification** |
| **SLE** | **Systemic lupus erythematosus** |
| **SOP** | **Standard operating procedure** |
| **SUSAR** | **Suspected unexpected serious adverse reaction** |
| **TCR** | **T cell receptor** |
| **TREC** | **T cell receptor excision circles** |
| **UAR** | **Unexpected adverse reaction** |
| **VFR** | **Vienna Food Record** |

#### Responsibilities and addresses

Sponsor:
Medical University Graz
Auenbruggerplatz 2
8036, Graz

Principal Investigator (in accordance with § 2a/Z11 and §35 of the Austrian Act on Medicinal Products):
**Assoz.Prof. PD Dr. med. univ. Martin Stradner**
Medical University Graz
Division of Rheumatology and Immunology
Auenbruggerplatz 15
8036 Graz – Austria
+43 316-385-81794


CO-PRINCIPAL INVESTIGATORS:

**Univ.-Prof. Dr. med. Ivo Steinmetz**
Medical University Graz
Institute of Hygiene, Microbiology and Environmental Medicine
Neue Stiftingtalstraße 6/III
8010 Graz – Austria
+43 316-385-73700


**Univ.-Prof. Dr. med. univ. Hildegard Greinix**
Medical University Graz
Division of Hematology
Auenbruggerplatz 38
8036 Graz – Austria
+43 316-385-81518


**Univ.-Prof. Dr. med. Peter Schlenke**
Medical University Graz
Department of Blood Group Serology and Transfusion Medicine
Auenbruggerplatz 48,
8036 Graz - Austria
+43 316-385-83067


<u>Study centre:</u>
Medical University Graz
Auenbruggerplatz 2
8036, Graz

<u>Biometry:</u>
**PD. Dr. med. Florian Posch**
Department of Oncology
Auenbruggerplatz 15
8036 Graz - Austria
+43 316-385-80173


<u>Biobanking</u>
**Monika Valjan, MA**
Biobank Graz
Neue Stiftingtalstraße 2
8010 Graz - Austria
+43 316-385-72719


#### Synopsis

| | |
|---|---|
| **Sponsor** | Medical University of Graz |
| **Name** | Humoral and cellular immune response to COVID-19 vaccines in immunocompromised and healthy individuals. |
| **Running head** | CoVVac study |
| **Target population (or indication)** | Patients with primary or secondary immunodeficiency planning on vaccination against SARS CoV-2 |
| **Study design and phase** | A prospective monocentric phase IV cohort study |
| **Version of protocol** | 3.0 |
| **Total duration of the trial** | 38 months |
| **Aims of the clinical trial** | <u>Primary aim of the trial</u><br><br>Characterization of the anti-SARS-CoV-2 humoral immune response after vaccination with a focus on the anti-spike protein response: an analysis in immunocompromised patients compared to healthy controls.<br><br><u>Secondary aims of the trial</u><br><br>To answer the following research questions:<br>1. Is there a difference in the T cell response between healthy subjects and immunodeficient patients?<br>2. Is there a difference in seroconversion or T cell response between different vaccines?<br>3. How does aging of the immune system impact the humoral and cellular immune response?<br>4. What is the impact of the various forms and severities of immunosuppression/immunodeficiency due to malignancy and/or immunosuppressive therapies on the vaccine-induced immune response? |

| | |
|---|---|
| | 5. What is the impact of a previous SARS-CoV-2 infection and its severity on the immune response after vaccination? e.g. the recognized epitopes/antigens, the distribution of immunoglobulin classes, the occurrence of neutralizing antibodies and the quantity and persistence of the vaccine response?<br>6. What is the role of previous asymptomatic (undiagnosed) SARS-CoV-2 infection (proven through specific antibodies) on the vaccine induced response?<br>7. How does the immune response differ between the various vaccines in immunocompromised individuals, COVID-19 recovered individuals and age-matched healthy controls?<br>8. Is there any difference in the induction of secretory IgA and serum IgA in comparison to the serum IgG and IgM response in immunocompromised individuals and in COVID-19 recovered individuals compared to controls?<br>9. What is the influence of previous infections caused by endemic CoV (proven through cross-reactive antibodies) on the vaccine response in immunocompromised individuals and in COVID-19 recovered individuals compared to controls?<br>10. Is there any difference in the induction of neutralizing antibodies in subsets of immunocompromised individuals and in COVID-19 recovered individuals compared to COVID-19 naïve controls?<br>11. Is there any difference in the neutralizing capacity of antibodies after vaccination towards the emerging SARS-CoV-2 variants in immunocompromised individuals and in COVID-19 recovered individuals compared to controls?<br>12. Do diet and body fat have an impact on the extent of seroconversion after vaccination? |
| **Outcome measures (endpoints) of the clinical trial** | <u>Primary outcome measure</u><br><br>The levels of anti-SARS-CoV-2 spike protein humoral immune response at day 21-28 after the second vaccination measured by SARS-CoV-2 |

| | |
|---|---|
| | antigen-binding Ig assay, comparing immunocompromised patients to healthy controls.<br>Secondary outcome measures<br>• Seroconversion measured by SARS-CoV-2 antigen-binding Ig assay 6, 12 and 24 months after vaccination<br>• Concentrations of recombinant S protein-binding IgG after second vaccination in comparison to response after first vaccination.<br>• Concentrations of recombinant S protein-binding IgG depending on prior exposure to SARS-CoV-2.<br>• Concentrations of secretory and serum IgA in comparison to IgG and IgM after second vaccination in immunocompromised, in recovered individuals and in healthy controls.<br>• IFNγ production of T cells after SARS-CoV-2 antigen exposure, measured by FACS and ELISpot.<br>• Identification of parameters predicting the response to COVID-19 vaccinations: prior CoV infection (cross-reactive antibodies), quantitative immunoglobulins, B cell subsets, T cell subsets; T cell aging (TCR diversity, telomere length, TREC levels).<br>• Neutralizing capacity of antibodies in respect of different SARS-CoV-2 variants.<br>• Correlation of dietary inflammation index and subcutaneous fat thickness sum with seroconversion after second vaccination. |
| **Number of patients** | 195 immunodeficient patients and 195 healthy controls |
| **Time schedule** | With reference to the trial<br>Recruitment period: March 2021 - December 2021<br>Planned start (FPFV): March 2021<br>Planned end (LPLV): May 2024<br>With reference to patients |

| | |
|---|---|
| | Individual duration of Study: 25 to 29 months |
| **Inclusion criteria** | 1. Noninfectious immunocompetent participants (healthy participants) as determined by medical history and clinical judgement.<br>or<br>2. Patients with primary immunodeficiencies<br>or<br>3. Patients with B-cell depleting therapy due autoimmune disease<br>or<br>4. Patients with benign and malignant hematological diseases receiving specific treatments with known immunosuppressive effects including cytotoxic agents, systemic corticosteroids, monoclonal antibodies and targeted therapies.<br>or<br>5. Patients with active hematological diseases and secondary immunoglobulin deficiency (e.g. CLL, MM) currently not receiving specific treatment.<br>or<br>6. Patients >3 months but <12 months after autologous HSCT.<br>or<br>7. Patients >3 months but <12 months after allogeneic HSCT.<br>or<br>8. Recipients of HSCT >12 months after allogeneic HSCT but under immunosuppressive therapy.<br>or<br>9. Patients with chronic GvHD and persistent immunodeficiency. |
| **Exclusion criteria** | *Healthy participants*<br>1. Presence of diseases or therapies that are likely to interfere with the immune response to vaccination.<br>2. Presence of a disease requiring change in therapy during 4 weeks before enrollment.<br>3. Any contraindications to the vaccine planned to receive as listed in the product characteristics. |

| | |
|---|---|
| | 4. Lack of willingness to undergo serial blood draws and attend follow-up appointments.<br>5. Women who are pregnant or breastfeeding.<br>6. Previous vaccination with any coronavirus vaccine.<br>7. Persons who are not willing to sign the informed consents (biobank informed consent and study specific informed consent)<br><br>*Immunodeficient participants*<br>1. Patients with hematological diseases within three months from B-cell-depleting immunotherapy (rituximab, ofatumumab, obinutuzumab, blinatumomab, CAR-T cells).<br>2. Patients with hematological malignancies in remission and >12 months after end of specific therapy.<br>3. Patients within three months from HSCT.<br>4. Any contraindications to the vaccine planned to receive as listed in the product characteristics.<br>5. Lack of willingness to undergo serial blood draws and attend follow-up appointments.<br>6. Women who are pregnant or breastfeeding.<br>7. Previous vaccination with any coronavirus vaccine (exception: if serum prior to vaccination is available from the biobank).<br>8. Patients who are not willing to sign the informed consents (biobank informed consent and study specific informed consent). |
| **Product characteristics** | Comirnaty:<br>https://www.ema.europa.eu/en/documents/product-information/comirnaty-epar-product-information_de.pdf<br>Moderna Covid-19 vaccine:<br>https://www.ema.europa.eu/en/documents/product-information/covid-19-vaccine-moderna-epar-product-information_de.pdf<br>AstraZeneca COVID-19 vaccine:<br>https://www.ema.europa.eu/en/documents/product-information/covid-19-vaccine-astrazeneca-epar-product-information_de.pdf |

#### 1. Scientific background

**SARS-CoV-2 immunity and vaccines**

The enveloped RNA virus SARS-CoV-2 and causative agent of the COVID-19 pandemic belongs to the family of *Coronaviridae*, which comprises a number of viruses that can cause disease in animals and humans. Among them are SARS-CoV and MERS-CoV and four endemic coronaviruses (HCoV-229E, -NL63, -OC43, and -HKU1) that cause relatively mild upper and lower respiratory tract infections in humans (1,2).

The induction of protective anti-SARS-CoV-2 immunity is a key strategy in the containment of the COVID-19 pandemic. Vaccine data from infection models with non-human primates indicate a correlation between the induction of neutralizing antibodies and reduced viral loads (3,4). The successful treatment of COVID-19 patients with convalescent plasma (5) and the recent emergency use authorization for neutralizing monoclonal antibodies by the United States Food and Drug Administration strongly support an important role for antibodies in anti-SARS-CoV-2 immunity (6). Furthermore, SARS-CoV-2 –specific CD4+ and CD8+ T cell responses in patients correlated with less severe disease suggesting a protective role in COVID-19 (7,8). Therefore, both, strong B cell and T cell mediated responses are likely to be necessary for an effective immunity.

Most studies have focused on the spike protein (S), containing the S1 and S2 subunits, as a vaccine target (9,10). The S1 subunit recognizes the host receptor human angiotensin converting enzyme 2 (ACE2) via its receptor-binding domain (RBD), whereas the S2 subunit is involved in membrane fusion and required for viral entry (11,12). The RBD is located at the C-terminal domain of the S1 subunit and is the major target for neutralizing antibodies, although the N-terminal domain is also recognized by neutralizing antibodies (13). The S2 subunit can also be recognized by antibodies with neutralizing capacity through interfering with membrane fusion (14). The SARS-CoV-2 structural proteins M (membrane) and E (envelope) seem to be less immunogenic but have a higher sequence homology with other corona viruses and exhibit cross-reactive T cell epitopes (15). The structural protein N (nucleocapsid) elicits a strong antibody response and also contains T cell epitopes (15), but has not been selected as a sole vaccine target against SARS-CoV-2 so far.

The S protein, either in full length with or without amino acid substitutions or as the S protein RBD, is the target of the two mRNA-based BNT162b2 (Pfizer-BioNTech) and mRNA-1273 (Moderna) and the vector-based vaccine ChAdOx1 nCoV-19 (AstraZeneca), which have recently been approved by the European Medicines Agency (EMA). These vaccines have been shown to induce humoral as well as cell mediated immune responses in the study populations (9,16,17). The efficacy of BNT162b2 to prevent COVID-19 was 95% (95% CI 90.3 to 97.6) with a similar efficacy in secondary analyses including age groups, ethnicity and body-mass index. Prevention of severe disease was also highly efficient (18). The overall efficacy of the mRNA-1273 vaccine was 94.1% (95% CI, 89.3 to 96.8). Again, secondary analyses revealed similar efficacies in subgroups stratified by age, sex, ethnicity and risk factors for severe COVID-19 (9). This vaccine was also shown to significantly reduce severe COVID-19. The efficacy of ChAdOx1 nCoV-19 reached 81.3% (95% CI 60·3–91·2) in an ongoing trial when the second dose was given after an interval of 12 or more weeks. More than 14 days after vaccination with the second dose, 15 COVID-19 hospitalizations occurred in the control group, but no hospitalization in the ChAdOx1 nCoV-19 group (19). A number of other vaccine candidates have entered phase III clinical trials and are expected to be available in the coming months (20).
The currently licensed vaccines induce a strong anti-S protein IgG response with virus neutralizing capacity (21–23). Available data on the anti-S protein T cell response of the mRNA-based vaccines demonstrate a predominant Th1-profile of SARS-CoV-2 specific CD4+ and CD8+ cells (22,23).

Since the vaccine efficacy trials did not include a comprehensive antibody status prior to vaccination (9,16,17), the impact of previous COVID-19 on the vaccine induced immune response, is unclear. Another significant gap of knowledge concerns the vaccine efficacy and immune response in severely immunocompromised individuals. Patients with acquired or innate immunodeficiencies were not included in the vaccine efficacy trials (9,16,17). Therefore, neither the type of induced immune response nor the efficacy of available vaccines in these patient cohorts have been thoroughly investigated.

So far, all available vaccine candidates are administered intramuscular or intradermal and the analysis of the corresponding antibody response has focused on serum antibodies. A gap of knowledge is the role of mucosal immunity in the defense against SARS-CoV-2, in particular the role of secretory immunoglobulins such as secretory IgA (SIgA). Although recent reports demonstrate that SARS-CoV-2-specific immunoglobulins can be detected in saliva after infection

(24) their persistence, diagnostic usefulness and protective capacity remains to be determined. In this context, it is unclear, if SARS-CoV-2-specific antibodies can be detected in mucosal secretions of the respiratory tract after immunization with the currently licensed vaccines and if a mucosal vaccine via the respiratory tract might further enhance immunity in this compartment.

The fact that mutations in the spike protein occur which not only alter the function of the protein but also the recognition by the immune system is of concern. Recently emerging SARS-CoV-2 variant strains from the UK, South Africa (SA) and Brazil appear to be more transmissible and accumulated a number of mutations/deletions in the spike protein that deserve close attention regarding the possible occurrence of vaccine escape variants (25). Currently most prevalent is the UK variant B.1.1.7, which is characterized by multiple changes in the spike protein including a deletion at position 69-70 and a mutation in the RBD at position 501. The latter resulting in a higher affinity to ACE2. The SA variant B.1.351 harbors multiple changes in the spike protein, with mutations K417N, E484K, and N501Y within the RBD, but no deletion 69-70. The Brazil variant P.1 accumulated also multiple amino acid changes in the spike protein, including K417T, E484K and N501Y in the RBD.

The efficacy of the vector-based vaccine ChAdOx1 nCoV-19 against the UK variant B.1.1.7 seems to be similar to the efficacy of the vaccine against other UK lineages (26). However, studies have shown that sera from either BNT162b2- or mRNA-1273-vaccinated individuals or convalescent plasma had a reduced neutralizing capacity towards engineered SARS CoV-2 variants with key spike mutations from the UK and SA variant (27,28). The neutralizing potency of a number of monoclonal antibodies recognizing different epitopes in the RBD were reduced or even abolished by either K417N, E484K, or N501Y mutations.

Taken together these results indicate the potential of SARS CoV-2 immune escape mechanisms after vaccination. It seems likely, that a continuous monitoring of the immunity of vaccinated individuals towards SARS CoV-2 variants will guide possible vaccine updates. This might be of particular relevance in patients with immunodeficiencies.

**SARS-CoV-2 and vaccination in patients with primary immunodeficiencies**

Immunodeficiency can be primary due to underlying genetic cause, like common variable immunodeficiency (CVID), or acquired (secondary immunodeficiencies), i.e., patients requiring immunosuppressive therapies due to autoimmune diseases (e.g. rheumatoid arthritis, SLE, vasculitis, IBD, multiple sclerosis), hematologic malignancies, after solid organ transplant or hematopoietic stem cell transplant. The ability to secrete protecting virus-specific antibodies and/or to create sufficient cellular immune responses is often reduced in these patients. Therefore, the question arises if they benefit from COVID-19 vaccines at all.

CVID is characterized by reduced immunoglobulin secretion and increased susceptibility to infection. In 2017, a review (29) already highlighted the immunization of CVID patients by influenza vaccination. It was demonstrated that the majority of tested CVID patients produced influenza-specific activated CD4+ T cells secreting IFN-γ, TNF-α and IL-2 on exposure to influenza antigens and, therefore, benefited from the vaccination despite their humoral immunodeficiency.

The immunogenicity of different vaccines has been examined in patients with secondary immunodeficiency due to rituximab and other antirheumatic therapies, as well. In a review from 2017, it was pointed out that influenza and pneumococcal vaccinations administered to patients taking B-cell depleting rituximab are significantly less immunogenic, i.e. an impaired humoral response occurs (30). In this context, however, dosing intervals played an important role: vaccinations given at least 6 months after last rituximab dose resulted in better humoral responses than short intervals of 1-2 months. A recent review about considerations of COVID-19 vaccines in patients with autoimmune diseases receiving anti-CD20-depleting therapies summarized that these patients do not necessarily suffer from severe SARS-CoV-2 infection because of the assumed central role of effective T cell responses in COVID-19 (31). Nevertheless, establishment of a protective immunity in terms of neutralizing antibodies following vaccination might be impaired until repopulation of naïve B cells (31), indicating that either intervals should be prolonged or that vaccination-induced cellular immunity could suffice, which needs further investigation now.

The potentially protective long-term effect of T cell responses to SARS-CoV-2 has recently been investigated in convalescents of the SARS outbreak in 2003 (32). Although 17 years ago, there still was a CD4+ and CD8+ T cell response to the N protein of SARS-CoV, which proved to be robustly cross-reactive against the N protein of SARS-CoV-2, as well. This could imply a lower susceptibility to COVID-19.

The special importance of cellular immune responses in primary immunodeficiencies was demonstrated in several cases of patients with X-linked agammaglobulinemia (XLA) and CVID who got infected with SARS-CoV-2 (33–35). Despite the defects in B cell differentiation and immunoglobulin production in these patients, they were able to recover – some even without need of oxygen ventilation, intensive care or treatment like COVID-19 convalescent plasma (34). These cases corroborate the important role of T cell responses in fighting COVID-19 (34).

**Vaccination and obesity**

An association between obesity and lower vaccine-induced H1N1-specific antibodies and obese individuals with double risk of developing influenza have been reported and the same is supposed for COVID-19 (36). However, it is still unclear whether obese adipose tissue may also alter vaccine response and vaccination side effects in patients with autoimmune deficiencies.

Diet and nutrient profile are crucial for the maintenance of health and appropriate immune function. Certain dietary components have the ability to shift the metabolic response and may trigger pro- and anti-inflammatory conditions (37). Thus, the evaluation of food and nutrient intake and the diet's inflammatory potential is indicated to assess the progression of the immune response to the vaccine.

**COVID-19 vaccination of patients with hematological malignancies**

Secondary immunodeficiencies in hematologic diseases can be caused by the malignant disease itself as in chronic lymphocytic leukemia (CLL), multiple myeloma (MM) and malignant lymphomas (NHL), its therapy (immunosuppression due to conventional chemotherapy, radiotherapy, targeted therapies, biological agents and immunotherapies) and/or associated metabolic dysfunctions (e.g. protein loss due to enteropathy, nephrotic syndrome with hypogammaglobulinemia, hepatic dysfunction, malnutrition with protein and vitamin deficiency). Furthermore, infections such as herpes viral infections can have a negative impact on patients' immune system. Secondary immunodeficiencies can affect both the innate as well as the adaptive immune system and increase patients' susceptibility for opportunistic infections that are one of the main causes of death of patients with hematologic malignancies (PHM). Immunodeficiency can also lead to reduced immune response towards leukemic or tumor cells. Patients with MM are uniquely susceptible to viral and bacterial diseases with a 7-to-10-fold higher risk of infection (38). MM and its treatments lead to humoral immunodeficiency, hypogammaglobulinemia and impaired B-and T-cell responses (39,40). MM usually affects the elderly population, a more vulnerable group of patients due to immunosenescence together with other comorbidities. Younger MM patients are frequently treated with high-dose chemotherapy followed by autologous hematopoietic stem cell transplantation (HSCT) with high infection susceptibility during the 3-month period following transplant (41). CLL is the most common form of leukemia and is associated with impaired immune responses to common pathogens in a context of profound immune dysregulation reflected by hypogammaglobulinemia, qualitative and quantitative B- and T-cell defects, including impaired response to vaccination, and CD4+ lymphopenia as well as innate immune dysfunction and neutropenia (42,43). In other diseases such as T-cell and Hodgkin lymphomas the clinically observed immunodeficiency has, so far, not been analyzed in more detail. Treatment of B-cell malignancies with anti-CD20 monoclonal antibodies such as rituximab causes profound B-cell depletion lasting in the majority of patients for a median of 6 to 9 months (44). The prevalence of hypogammaglobulinemia after rituximab therapy varies dependent on malignant disease and concomitant therapies and reportedly was 38% in a cohort of B-cell lymphoma patients including 6% with need for immunoglobulin substitution.

PHM have a high risk of developing severe COVID-19 disease course after SARS-CoV-2 infection. The first case study specifically examining PHM who became infected at the onset of the COVID-19 pandemic in Wuhan showed a high mortality of 38.5% (45). Further retrospective studies in other European centers confirmed this high fatality rate for PHM. A case series of 59 patients from four hospitals in Italy, Spain and the Netherlands showed an age dependent mortality rate of 34% (46). In Italy, data from 66 hospitals on 536 PHM with COVID-19 disease demonstrated a worse overall survival (OS) compared to PHM without COVID-19. Older age (hazard ratio [HR] 1.03, 95% CI 1.01–1.05), progression of malignant disease (HR 2.10, 95% CI 1.41–3.12), diagnosis of acute myeloid leukemia (HR 3,49, 95% CI 1,56–7,81), indolent NHL (HR 2.19, 95% CI 1.07–4.48), aggressive NHL (HR 2.56, 95% CI 1.34–4.89), or plasma cell neoplasms (HR 2.48, 95% CI 1.31–4.69) were associated with worse OS (47). A recent meta-analysis of 3377 PHM with COVID-19 demonstrated a case fatality rate of 34% (48). The pooled risk of death for several diseases was as listed: Acquired bone marrow failure syndromes (14 studies, 231 patients), 53% (95% CI, 34-72); acute leukemias (18 studies, 289 patients), 41% (95% CI, 30-52); plasma cell dyscrasias (23 studies, 412 patients), 33% (95% CI, 25-41); NHL, including CLL (20 studies, 1324 patients), 32% (95% CI, 24-40); NHL, excluding CLL (14 studies, 485 patients), 32% (95% CI, 18-48); CLL specifically (15 studies, 517 patients), 31% (95% CI, 23-40); myeloproliferative neoplasms (MPNs) (12 studies, 293 patients), 34% (95% CI, 19-51) (48). It must be noted that the majority (77%) of patients analyzed had been hospitalized.

PHM who received a hematopoietic stem-cell transplantation (HSCT) constitute an important group of immunodeficient individuals. After allogeneic HSCT patients reconstitute their immune system from donor-derived immune precursor cells in the stem cell graft. Complete immune regeneration can take 1 to 3 years depending on duration of immunosuppression for graft-versus-host disease (GvHD) prophylaxis and therapy. Several factors significantly influence immunologic reconstitution after allogeneic HSCT including the restoration of protective, anti-infective host immunity and the reestablishment of expanded T cell and B cell repertoires as well as functional tolerance with preservation of graft-versus-leukemia (GvL) effects. These factors include conditioning regimen intensity, graft source, donor type, HLA-identity, donor and recipient age (49). Immune dysregulation and alloreactivity drive the development of chronic GvHD resulting from the predominance of donor-derived effector mechanisms that cannot be controlled by donor- or host-derived regulatory immune responses. GvHD can attack the thymus resulting in the generation of donor T cells with antihost reactivity due to defective negative selection (50). The pathophysiology of chronic GvHD is complex and involves multiple distinct interactions among alloreactive and dysregulated T and B cells and innate immune cell populations (49). Patients with chronic GvHD can present with autoimmune phenomena, autoantibody production, severe immunodeficiency due to lack of T and B cells and functional asplenia.

The Center for International Blood and Marrow Transplant Research (CIBMTR) analyzed the outcome of 318 HSCT recipients who developed COVID-19. The estimated OS at 30 days after COVID-19 diagnosis was 68% (95% CI 58–77) for recipients of allogeneic HSCT and 67% (55–78) for autograft recipients (51). Male sex, age older than 50 years and being within a year from allogeneic HSCT were factors strongly associated with worse OS (51). These reported outcomes are comparable to results from other studies from two North American cohorts (52,53).

Vaccination is important in PHM because they have an increased risk of infection related morbidity and mortality due to common pathogens like influenza, pneumococci and varicella zoster virus. The new COVID-19 vaccines have not been tested in PHM or recipients of HSCT. It is unknown what level of protection and type of immune response will result from COVID-19 vaccination.
Most data on serologic response to COVID-19 infection have come from immunocompetent adults (54). In a small case series, nine of ten patients with acute leukemia on systemic anticancer therapies were able to induce anti-SARS-CoV-2 IgG antibody responses but only 67% of 30 patients with CLL (55,56).
Since the method of vaccination differs according to nature of malignancy, treatment, infection, and vaccine, the European Conference on Infections in Leukemia (ECIL 7) developed guidelines for PHM including HSCT recipients in 2017 (57,58). Most of these recommendations are not based on clinical efficacy but on serological responses because hematological malignancies are rare and the overall incidence of infections is low. Studies whose results were used to establish the ECIL7 guidelines show lower overall effectiveness of most vaccines (summarized by (57,58)). Of note, immunotherapy with rituximab at the time of vaccination (and probably other B-cell depleting

antibodies), strongly impairs vaccination response for at least 6 months. Since PHM have a high risk of poor outcome after COVID-19, the German Society of Hematology and Oncology recommends COVID-19 vaccination at the earliest 3 months after B-cell depleting therapy (59).

#### 2. Aim of the trial

Currently, the efficacy of COVID-19 vaccination in immunodeficient patients is unknown. Here we aim to evaluate the efficacy of COVID-19 vaccines in immunodeficient patients compared to healthy controls. We will assess the humoral and cellular response to COVID-19 vaccination in these subjects in detail. Furthermore, we will identify factors associated with good response to vaccination. The results of our study will help to guide future recommendations on COVID-19 vaccination in this population.

##### Primary objective

Characterization of the anti-SARS-CoV-2 humoral immune response after vaccination with a focus on the anti-spike protein response: an analysis in immunocompromised patients compared to healthy controls.

##### Secondary objectives

To answer the following research questions:

1. Is there a difference in the T cell response between healthy subjects and immunodeficient patients?
2. Is there a difference in seroconversion or T cell response between different vaccines?
3. How does aging of the immune system impact the humoral and cellular immune response?
4. What is the impact of the various forms and severities of immunosuppression/immunodeficiency due to malignancy and/or immunosuppressive therapies on the vaccine-induced immune response?
5. What is the impact of a previous SARS-CoV-2 infection on the immune response after vaccination? e.g. the recognized epitopes/antigens, the distribution of immunoglobulin classes, the occurrence of neutralizing antibodies and the quantity and persistence of the vaccine response?
6. What is the role of previous asymptomatic (undiagnosed) SARS-CoV-2 infection (proven through specific antibodies) on the vaccine induced response?

7. How does the immune response differ between the various vaccines in immunocompromised individuals, COVID-19 recovered individuals and age-matched healthy controls?
8. Is there any difference in the induction of secretory IgA and serum IgA in comparison to the serum IgG and IgM response in immunocompromised individuals and in COVID-19 recovered individuals compared to controls?
9. What is the influence of previous infections caused by endemic CoV (proven through cross-reactive antibodies) on the vaccine response in immunocompromised individuals and in COVID-19 recovered individuals compared to controls?
10. Is there any difference in the induction of neutralizing antibodies in subsets of immunocompromised individuals and in COVID-19 recovered individuals compared to COVID-19 naïve controls?
11. Is there any difference in the neutralizing capacity of antibodies after vaccination towards the emerging SARS-CoV-2 variants in immunocompromised individuals and in COVID-19 recovered individuals compared to controls?
12. Do diet and body fat have an impact on the extent of seroconversion after vaccination?

#### *2.1. Study design*

We will conduct a prospective cohort study including 195 adult participants with primary or secondary immunodeficiencies and 195 healthy control subjects at a single center. The study will take place at the Medical University of Graz. Blood and saliva samples will be obtained before COVID-19 vaccination (study visit 1), and after the second vaccination vaccinations (study visits 2-5, see Table 1).

##### 2.1.1 Time schedule

The total duration of the study is 38 months, starting in March 2021 with a recruiting period until 31. December 2021 and finish of all scheduled visits until 31. May 2024. Recruitment of patients and healthy controls will be organized as described under 3.1. Five on-site visits and one telephone visit are scheduled within an individual study period of 25 to 29 months. A detailed description of the time schedule and the individual visits is provided under 4.

#### *2.2 Outcome measures*

##### 2.2.1 Primary outcome measures

Levels of anti-SARS-CoV-2 spike protein antibodies at day 21-28 after the second vaccination measured by SARS-CoV-2 antigen-binding Ig assay.

##### 2.2.2 Secondary outcome measures

- Seroconversion measured by SARS-CoV-2 antigen-binding Ig assay 6, 12 and 24 months after vaccination
- Concentrations of recombinant S protein-binding IgG after second vaccination in comparison to response after first vaccination.
- Concentrations of recombinant S protein-binding IgG depending on prior exposure to SARS-CoV-2.
- Concentrations of secretory and serum IgA in comparison to IgG and IgM after second vaccination in immunocompromised, in recovered individuals and in healthy controls.
- IFNγ production of T cells after SARS-CoV-2 antigen exposure, measured by FACS and ELISpot.
- Identification of parameters predicting the response to COVID-19 vaccinations: prior CoV infection (cross-reactive antibodies), quantitative immunoglobulins, B cell subsets, T cell subsets; T cell aging (TCR diversity, telomere length, TREC levels).
- Neutralizing capacity of antibodies in respect of different SARS-CoV-2 variants.
- Correlation of dietary inflammation index and subcutaneous fat thickness sum with seroconversion after second vaccination.

### 3. Recruitment of probands

#### *3.1 Number of probands and the duration of the trial*

Male and female subjects older than 18 years will be recruited before undergoing COVID-19 vaccination according to the Austrian vaccination plan. 195 immunodeficient patients will be recruited at the Medical University of Graz (Division of Rheumatology and Immunology, Division of Hematology). 195 healthy controls will be recruited from 1.) already existing Covid-19 convalescent cohort (Department of Internal Medicine, Section of Infectious Diseases and Tropical Medicine, Department of Blood Group Serology and Transfusion Medicine, Institute of Hygiene, Microbiology and Environmental Medicine); 2.) healthy controls such as whole blood and platelet donors (Department of Blood Group Serology and Transfusion Medicine) or other volunteers recruited by adequate advertisement. Vaccination of immunodeficient patients will occur prior to healthy controls according to the Austrian vaccination plan. Therefore, healthy controls will be selected to match immunodeficient patients' age, sex and type of vaccine and will be recruited as soon as the immunodeficient group is characterized and according to the Austrian vaccination plan, i.e. prospectively from June 2021 on. The individual duration of the study will be 25 to 29 months depending on the type of vaccine.

##### *3.2 Inclusion criteria*

In order to be enrolled, participants must be 18 years or older, able to understand study procedures, provide written informed consent (Biobank informed consent and study specific informed consent), and meet one of the following inclusion criteria:

1. Noninfectious immunocompetent participants (i.e., healthy participants) as determined by medical history and clinical judgement.

   or
2. Patients with primary immunodeficiencies

   or
3. Patients with B-cell depleting therapy due autoimmune disease

   or
4. Patients with benign and malignant hematological diseases receiving specific treatments with known immunosuppressive effects including cytotoxic agents, systemic corticosteroids, monoclonal antibodies and targeted therapies.

   or
5. Patients with active hematological diseases and secondary immunoglobulin deficiency (e.g. CLL, MM) currently not receiving specific treatment.

   or
6. Patients >3 months but <12 months after autologous HSCT.

or

7. Patients >3 months but <12 months after allogeneic HSCT.

or

8. Recipients of HSCT >12 months after allogeneic HSCT but under immunosuppressive therapy.

or

9. Patients with chronic GvHD and persistent immunodeficiency.

##### *3.3 Exclusion criteria*

Subjects meeting any of the following criteria cannot be enrolled into the trial:

*Healthy participants*

1. Presence of diseases or therapies that are likely to interfere with the immune response to vaccination.
2. Presence of a disease requiring change in therapy during 4 weeks before enrollment.
3. Any contraindications to the vaccine planned to receive as listed in the product characteristics.
4. Lack of willingness to undergo serial blood draws and attend follow-up appointments.
5. Women who are pregnant or breastfeeding.
6. Previous vaccination with any coronavirus vaccine.
7. Persons who are not willing to sign the informed consents (biobank informed consent and study specific informed consent).

*Immunodeficient participants*

1. Patients with hematological diseases within three months from B-cell-depleting immunotherapy (rituximab, ofatumumab, obinutuzumab, blinatumomab, CAR-T cells).
2. Patients with hematological malignancies in remission and >12 months after end of specific therapy.
3. Patients within three months from HSCT.
4. Any contraindications to the vaccine planned to receive as listed in the product characteristics.
5. Lack of willingness to undergo serial blood draws and attend follow-up appointments.

6. Women who are pregnant or breastfeeding.
7. Previous vaccination with any coronavirus vaccine (exception: if serum prior to vaccination is available from the biobank).
8. Patients who are not willing to sign the informed consents (biobank informed consent and study specific informed consent).

##### 3.4 Clinical history

Date and type of COVID-19 vaccinations, medical history (including immunosuppressive therapies, chronic GvHD, severe opportunistic infections, prior vaccination history (types, titers if available), current and past disease including COVID-19), current medication, BMI, smoking habits, and nutritive assessment will be obtained in all participants.

##### 3.5 Parameters, Laboratory parameters

In order to assess parameters predicting the vaccination response, quantitative immunoglobulins and B cell subsets will be measured at the Diagnostic Immunology Lab of the Div. of Rheumatology and Immunology at baseline (visit 1). Pregnancy tests will be performed in the urine of female participants by detecting Human chorionic gonadotropin beta (once before each vaccination).

#### 4. Implementation of the trial / Course of the trial

The total duration of the study is 38 months, starting in March 2021 with a recruiting period until 31. December 2021 and finish of all scheduled visits until 31. May 2024. Recruitment of patients and healthy controls will be organized as described under 3.1. Individuals having consented and fulfilling the inclusion and exclusion criteria are included in the trial. A baseline visit will take place up to 60 days before the planned date of vaccination according to the Austrian vaccination plan. Scheduled vaccination with any COVID-19 vaccine approved in Austria will allow recruitment. If appropriate pre-vaccine samples from study participants exist in the biobank of the Medical University of Graz, patients may also be included in the study after vaccination starting with visit 3. In this case the biomaterial available at the biobank will be used for the analyses planned on visit 1. The investigators will not influence the date of vaccination or the type of the vaccine used. After first vaccination a telephone visit (visit 2) will assess adverse events and schedule visit 3 at the appropriate time after the second vaccination. At visit 3 the patient`s vaccination certificate will be checked to verify correct vaccination and document the type of vaccine received. Further visits (5-6) will be performed for up to two years after the second vaccination. This follow-up period will allow an assessment of the duration of the immune response. A detailed description of the individual visits is provided under 4.1 and in table 1. The data recorded directly on the CRF are considered to be source data. COVID-19 vaccination, COVID-19 infection, Vaccination history, Pregnancy test, Adverse Events, BMI, and Laboratory Specimen Collection are recorded directly on the CRF and therefore are considered to be source data. Data will be merged in an electronic database (RDA Research, Documentation & Analysis; Medical University of Graz, version 07.03.2019). CRFs will be inspected in respect of their accuracy and completeness and compared to original data by the monitor.

##### *4.1 Description of the individual visits*

**Visit 1**:
After informed consents, inclusion and exclusion criteria will be checked to include or exclude the subject in the trial. Clinical history (see 3.4) will be assessed and blood (107ml – Serology, immune status, T cell immunity, and T cell aging) and saliva (1.5ml) will be taken. Urine from female participants will be collected to perform a pregnancy test. The subjects will be provided with the Vienna Food Record questionnaire and one further pregnancy test for use before 2nd vaccination.
**Visit 2**:
Visit 2 (0-14 days after first vaccination) is a telephone visit assessing safety and scheduling visit 3 at the appropriate time after the second vaccination. Female participants are reminded to perform a pregnancy test in urine before receiving the 2nd vaccination.
**Visit 3**:
21-28 days after 2nd vaccination, vaccination reactions and clinical history will be assessed. Blood (68ml – serology, T cell immunity) and saliva (1.5ml) will be taken. Safety monitoring will be performed. The Vienna Food Record questionnaire will be collected and body fat composition will be measured by ultra sound.
**Visit 4**:
6 Month (±3 weeks) after 2nd vaccination, clinical history will be assessed. Blood (12ml – serology, blood group) and saliva (1.5ml) will be taken. Safety monitoring will be performed. The subjects will be provided with the Vienna Food Record questionnaire.
**Visit 5**:
12 Month (±3 weeks) after 2nd vaccination, clinical history will be assessed. Blood (68ml – serology, T cell immunity) and saliva (1.5ml) will be taken. Safety monitoring will be performed. The Vienna Food Record questionnaire will be collected and body fat composition will be measured by ultra sound.
**Visit 6**:
24 Month (±3 weeks) after 2nd vaccination, clinical history will be assessed. Blood (68ml – serology, T cell immunity) and saliva (1.5ml) will be taken. Safety monitoring will be performed.

| Visits | 1 | 2 | 3 | 4 | 5 | 6 |
|---|---|---|---|---|---|---|
| Time points | *60-0 days before vaccination* | *0-14 days after first vaccination* | *+ 21-28 days after 2nd vaccination* | *+6 months after 2nd vaccination* | *+12 months after 2nd vaccination* | *+24 months after 2nd vaccination* |
| Informed consent | X | | (x) | | | |
| Inclusion/Exclusion criteria | X | | | | | |
| Medical history incl. COVID-19 | X | | X | X | X | X |
| Telephone visit | | X | | | | |
| Serology + saliva | X | | X | X | X | X |
| T cell immunity | X | | X | | X | X |
| T cell aging | X | | | | | |
| Immune status | X | | | | | |
| Nutritive assessment | | | X | | X | |
| Body fat composition | | | X | | X | |
| Pregnancy test | X | (X) | | | | |
| Safety monitoring | | X | X | X | X | X |

**Table 1.** Visits and assessment schedule

##### *4.2 Methods*

**Sampling**

For serology, 8 ml of blood and 1.5 ml saliva samples will be taken at every visit.

For testing cellular immunity, additional 60 ml of blood will be drawn at visits 1, 3, 5 and 6. Assessment of T cell aging requires 20 ml of blood at visit 1. In order to assess parameters predicting the vaccination response (immune status) 19ml of blood will be drawn at visit 1. Blood group antigens (3 ml) will be determined at visit 4. Pregnancy tests will be performed in the urine of female participants by detecting Human chorionic gonadotropin beta.

**Storage**

All samples will be stored at the Biobank of the Medical University of Graz for further analysis. Samples that are not consumed by analyses of the current study will be made available for further

serologic, immunologic, genetic, or metabolomics analyses. For further analyses, not covered by this protocol, a separate ethical vote has to be obtained.
Serum samples will be stored at -80°C. Peripheral blood mononucleated cells will be frozen in DMSO-based media and stored in liquid nitrogen. Saliva will be collected by using the "passive drool" method. Alternatively, specimens will be collected using a device which is placed into the oral cavity or by a mouth wash. Details of the procedures are provided in the respective SOPs. Handling and storage of saliva samples will take place at the Institute of Hygiene, Microbiology and Environmental Medicine. In order to remove cell debris, the samples will be centrifuged for 15 min with 10.000g at 4°C and the supernatant will be stored at -80°C with constant temperature monitoring.

**Antibody tests**

Antibody tests with serum and saliva specimens will be conducted at the R&D Institute of Hygiene, Microbiology and Environmental Medicine. Anti-SARS-CoV-2 Ig isotypes against various forms of the S and N protein as well as the Ig-mediated immune response against other viral and bacterial antigens will be determined by using immunoassays currently applied in routine diagnostics. SARS-CoV-2 Live virus neutralization assays will be conducted in our biosafety level 3 laboratory. Either commercially available recombinant SARS-CoV-2 wild type proteins, homologous proteins of related viruses and SARS-CoV-2 variant proteins or recombinant SARS-CoV-2 proteins produced in our laboratory will be used in multiplex immunoassays for an in-depth analysis of the humoral immune response.

**T cell assays**

As part of the clinical studies for approval of the COVID-19 vaccines, virus-specific CD4+ and CD8+ T cell responses were investigated by IFNγ enzyme-linked immunosorbent spot (ELISpot) (22). In this study, we will perform ELISpot and other methods for research on T cell responses, such as fluorescence-activated cell sorting (FACS), before and after COVID-19 vaccination. Moreover, we will examine T cell aging in our patients as possible pre-existing factor for diminished immune responses to vaccinations.

*Isolation of PBMCs & T cell subsets*

As a precondition for all further T cell analyses explained below, peripheral blood mononuclear cells (PBMCs) will be isolated by Ficoll density gradient centrifugation. Total or naïve CD4+ and CD8+ T cells will be obtained using MACS technology according to manufacturer's instructions.

*Immunophenotyping and flow cytometry*

In order to detect and quantify SARS-CoV-2-specific T cells by stimulation of their cytokine secretion, flow cytometry and ELISpot will be performed.

Immunophenotyping will be executed on PBMCs directly ex vivo. Cytokine-producing T cells will be identified by intracellular cytokine staining. For that reason, PBMCs will be restimulated with a peptide pool representing the vaccine-encoded SARS-CoV-2 RBD (2 µg/ml/peptide; JPT Peptide Technologies) in the presence of GolgiPlug (BD Biosciences, San Jose, CA, USA) for 18 h at 37 °C. For both approaches, surface staining will be performed for 20 min at 4 °C. Afterwards, samples will be fixed and permeabilized using the Cytofix/Cytoperm kit according to the manufacturer's instructions (BD Biosciences). Intracellular staining will be performed in Perm/Wash buffer for 30 min at 4 °C. Samples will be acquired on a FACSLyric instrument (BD Biosciences) analyzed with FlowJo software version 10.5.3 (FlowJo LLC).

*ELISpot*

IFNγ ELISpot analysis will be performed using 3.3 × 105 isolated CD4+ and CD8+ T cells per well according to the manufactorer's instructions. In brief, cells will be stimulated for 16–20 h with an overlapping peptide pool representing the vaccine-encoded RBD. Bound IFNγ will be visualized using a secondary anti-IFNγ antibody directly conjugated with alkaline phosphatase (1:250; ELISpotPro kit, Mabtech AB, Nacka Strand, Sweden) followed by incubation with a 5-bromo-4-chloro-3'-indolyl phosphate (BCIP)/nitro blue tetrazolium (NBT) substrate (ELISpotPro kit, Mabtech). Plates will be scanned using an AID Classic Robot ELISPOT Reader and analyzed by AID ELISPOT 7.0 software (AID Autoimmun Diagnostika, Straßberg, Germany).

*Cytokine measurement*

To detect a variety of different cytokines produced by T cells after exposure to the S protein, additional Cytokine measurements will be done. PBMCs will be restimulated for 48 h with SARS-CoV-2 RBD peptide pool (2 µg/ml). Concentrations of various cytokines in supernatants will be determined using a magnetic bead-based Multiplex technology (MagPix, Merc, Darmstadt, Germany) according to the manufacturer's instructions.

**Methods on T cell aging**

*TCR diversity*

RNA will be extracted using RNeasy Protect Mini Kit (Qiagen, Germantown, MD, USA) according to the manufacturer instructions. RNA will be used for reverse transcription with First Strand cDNA

Synthesis Kit for RT-PCR AMV (Roche) according to the manufacturer regulations. cDNA will be diluted 1:5 for PCR using AmpliTaq Gold™ DNA Polymerase (Applied Biosystems, Waltham, MA, USA), 1× PCR Gold Buffer (Applied Biosystems), 2.5mM MgCl2 (Applied Biosystems), 0.4 mM dNTP Polymerization Mix (GE Healthcare), 0.5 µM TCR C β 5′FAM labeled primer (Ingenetix, Vienna, Austria), and 0.5 µM unlabeled TCR V β primers (Ingenetix). Cycle conditions will be a denaturation step at 94°C for 6 min, 35 cycles of 94°C for 1 min, 59°C for 1 min, and 72°C for 1 min, a final annealing step at 72°C for 7 min. After amplification, the PCR product will be supplemented with GeneSCan™-500 TAMRA™ Size Standard (Applied Biosystems) and HI-DI Formamide (Applied Biosystems). Electrophoresis will be performed with ABI Prism 310 Genetic Analyzer (Applied Biosystems), and 310 Data Collection Software. Analysis will be done by GeneScan® Software (Applied Biosystems). Calculations will include peak count (Complexity score) and single peak area in percent of whole peak area.

*Telomere length analysis*

We will measure telomere lengths using DNA from isolated naïve CD4+ and CD8+ T cell subsets by quantitative real-time PCR analysis and LightCycler FastStart DNA Master SYBR Green I (Roche) according to manufacturer's instructions. The following primers will be used: 5´-CGGTTTGTTTGGGTTTGGGTTTGGGTTTGGGTTTGGGTT-3´ and 5´GGCTTGCCTTACCCTTACCCTTACCCTTACCCTTACCCT-3´. A single copy gene (36B4) (fw primer: 5´CCCATTCTATCATCAACGGGTACAA-3´; rev primer: 5´CAGCAAGTGGGAAGGTGTAATCC-3´) served for normalization. Ct-values of test samples and control single copy genes will be calculated and 2ΔCt value will be determined.

*TREC measurement*

T cell receptor excision circles (TRECs) from naïve CD4+ and CD8+ T cells will be analyzed. In brief, DNA will be extracted using QIAamp DNA Mini Kit (Qiagen, Chatsworth, CA, USA). TREC concentrations will be determined by quantitative real-time PCR based on the coding TREC sequence using LightCycler 480 II (Roche) and primers to amplify a DNA fragment 108 bp across the remaining recombination sequence δrec/ψalpha (5'-CACATCCCTTTCAACCATGCT-3' and 5'-GGTGCAGGTGCCTATGC-3'). For quantification, a synthetic internal standard (Ingenetix, Vienna, Austria) will be used. PCR reaction will run with 0.25 µg DNA, primers and hybridisation-probe labelled with FAM-TAMRA (5'-ACACCTCTGGTTTTTGTA AAGGTGCCCAC-3'). To avoid bias by different numbers of naïve T cells, TRECs will be calculated in relation to numbers of T cells.

**Nutritive assessment**

*Evaluation of Nutrient and Energy Intake:*

To assess the nutrient and energy intake as well as the participants' dietary habits an expanded version of the Vienna Food Record (VFR) will be used (60). The VFR has been developed in accordance with Austrian eating preferences and habits, considers commonly available food and is weighted for consumption frequencies in Austria based on data of the European Food Safety Authority (EFSA). It is a prospective food frequency questionnaire that enables insides into the individual's dietary patterns and nutrient and energy intake. The participants will be asked to report their food intake of four days including one weekend day prior the second vaccination.

The diet's nutrient and energy composition will be analysed by a national specific nutritional software (dato Denkwerkzeuge 2021) that comprises an Austrian specific food and nutrient database.

*Calculation of the Dietary Inflammation Index:*

Since diet plays an important role in the regulation of chronic inflammation, the dietary inflammation index (DII) will be applied to the food records. The DII is a tool that categorizes the individual's diet on a continuum from maximally anti-inflammatory to maximally pro-inflammatory. The DII can be used as a categorical variable, and predicts the overall inflammatory potential of the diet in accordance with a set of global norms (37,61). The DII will be calculated based on the VFR data.

*Measurement of Body Fat and Fat Patterning:*

For the determination of the patient's body fat and the fat patterning the ultrasound method will be applied (62,63). This method reaches an extremely high accuracy and is capable of detecting subcutaneous adipose tissue thicknesses on a fine scale in people of various body composition (64). The thickness of subcutaneous fat will be measured at eight precisely defined body sites and the thickness sum will be calculated ($D_{incl}$). Anthropometric measurements of body mass (m), body height (h), sitting height (s) and the circumferences of waist and hip will be performed in accordance to the International Society for the Advancement of Kinanthropometry (ISAK) (65).

##### *4.3 Risk-benefit assessment and precautionary measures*

Risk from blood draws and collection of biomaterial and data is minimal. Participants of the study will benefit by knowing the levels of neutralizing antibodies to SARS CoV-2, the main factor for sterilizing immunity. Furthermore, our study will add to the knowledge on vaccination in immunodeficient patients and will inform future guidelines. The trial is carried out according to GCP and legal principles as outlined under 8.

###### 4.3.1 Pregnancy tests

Pregnancy tests will be conducted before each vaccination. In the inactive phase of the study (visits 3-6), no increased study-related risk can be expected for pregnant women or unborn children. Therefore, no further pregnancy tests are planned in this phase.

##### *4.4 Adverse events*

Scheduled vaccination with any COVID-19 vaccine approved in Austria will not take place in the course of the trial but will happen according to the Austrian vaccination schedule. The investigators will not influence the date of vaccination or the type of the vaccine used. Adverse Events related to COVID-19 vaccines might occur.

Furthermore, events related to study procedures blood draw and salvia collection might occur.

Based on these facts, safety monitoring will be performed as follows:

- Any Adverse Event related to COVID-19 vaccines,
- Any Adverse Event related to blood draw and salvia collection,
- Any Serious Adverse Event.

Safety monitoring will be performed at every visit.

###### 4.4.1 Definitions

An **adverse event (AE)** is defined as any harmful incident experienced by a study participant, not necessarily causally related to the clinical trial.

An **adverse reaction (AR)** is any harmful unintentional reaction to a study medication.

An **unexpected adverse reaction (UAR)** is an adverse effect which, in terms of its type and severity, is not anticipated on the basis of the existing product information.
A **serious adverse event (SAE) or a serious adverse reaction (SAR)** is an adverse event or a serious adverse effect that is fatal or life-threatening (independent of the dose), necessitates in-hospital treatment or its prolongation, leads to permanent or serious disability or incapacity or a congenital abnormality or birth defect.
A **suspected unexpected serious adverse reaction (SUSAR)** is designated as such according to Guideline 2001/20/EG. A serious adverse reaction is deemed unexpected when it is not listed in the corresponding basic document (Summary of Product Characteristics, IB, IMPD).

##### 4.4.2 Reporting obligations

Principal Investigator:
Reporting AE as specified above, SAE and SUSAR to the sponsor according to AMG (Austrian Act on Pharmaceutical Products) § 41d.

Sponsor:
The sponsor must report all important information about suspected serious adverse to the Federal Agency for Safety in Healthcare as well as the competent ethics committee according to AMG §41e (Austrian Act on Pharmaceutical Products §41e).

##### *4.5 Interim analyses*

An interim analysis will be done when at least 30% of the subjects have completed visit 3. At that point, we will check if the described methods have worked properly and provided valid, comparable results. Interim analysis will predominantly include the primary endpoint and also the secondary endpoints of T cell responses. Depending on the results, the protocol may be amended. A change in sample size based on the results of the interim analysis will also lead to an amendment of the protocol. Amendments will be reported to the ethics committee and authorities as detailed under 7.

##### *4.6 Termination of the trial*

###### 4.6.1 Termination of the trial for a subject (drop-out)

Participants may discontinue study procedures and withdraw from the study at his/her own request at any time. Definitive discontinuation is also possible in case of adverse events after the first dose of vaccination, request of the investigator or diagnosis of pregnancy. Violation of the protocol or incorrect vaccination, as documented in the vaccination certificate will lead to termination of the trial for the subject. In case of discontinuation, participants will be followed for safety, and already collected data as well as biomaterial will still be analyzed according to study protocol. After withdrawal of consent, participants may demand that collected subject data have to be deleted and biomaterial must be destroyed. The investigator must document the reason of discontinuation or withdrawal and any request made by participants.

###### 4.6.2 Termination of the entire trial

Premature termination of the clinical trial will be considered when the sponsor believes it is necessary to terminate the clinical trial for safety reasons, or when the clinical trial proves to be impracticable.

For the benefit of, and in the interest of the probands, the Principal Investigator may terminate the trial at any time when serious adverse effects or other unforeseen circumstances occur.

Reporting obligation in case of termination: In case of termination of the trial, the responsible ethics committee and the authorities (BASG) must be informed within 15 days; the reasons for termination must be clearly stated herein.

#### 5. Monitoring and Audit – Assurance of data quality

Monitoring and audits shall be performed during the clinical trial for the purpose of quality assurance.

##### 5.1 Monitoring

The investigator consents to data evaluation being performed by the person in charge of monitoring in order to ensure satisfactory data collection and adherence to the study protocol. Furthermore, the investigator states that he/she is willing to cooperate with this person and shall provide this person with all required information whenever necessary. This includes access to all documents associated with the trial, including study-relevant medical files of patients in original form. The tasks of the investigator include maintenance of these patients' medical files as comprehensively as possible; this includes information concerning medical history, accompanying diseases, inclusion in the trial, data about visits, results of investigations, dispensing medication, and adverse events. The monitor will also be permitted to perform data evaluation and draw comparisons with the relevant medical files in accordance with the SOPs and ICH-GCP guidelines at pre-determined intervals, in order to ensure adherence to the study protocol and continuous registration of data. All original medical reports required as sources for the information given in the CRF or the database shall be inspected. The study participants will have given their consent to such inspection by signing the consent form.
The person in charge of monitoring is obliged to treat all information as confidential, and to preserve the basic claims of the study participants in respect of integrity and protection of their privacy.

The purpose of these visits specifically includes the following:

- Inspection of informed consent forms,
- Inspection of patient safety (occurrence and documentation / reporting of AEs and SAEs),
- Inspection of CRFs in respect of their accuracy and completeness,
- Validation of CRFs compared to original data (source data verification, SDV),
- Evaluation of the progress of the trial,
- Control of compliance with the study protocol,
- Ensuring that the trial is being performed at the study centre in conformity with GCP
- Discussion with the principal investigator about the implementation of the trial and any deficiencies that may have been noted

A monitoring report shall be issued about every visit. The report will document the progress of the clinical trial and will provide information about any difficulties that may have occurred.

##### *5.2 Audit*

The sponsor is authorised to perform audits at the study centre and other facilities that may be involved in the trial, as part of quality assurance. The purpose of the audit is to check the validity, verifiability, and completeness of the data and the credibility of the clinical trial as well as inspect the maintenance of patients' rights and patient safety. The sponsor may authorise persons not otherwise involved in the clinical trial for this task (auditors). These persons shall be permitted to inspect all study-related documents (especially the study protocol, survey forms, medical files, documentation of the study medication, study-related correspondence).
The sponsor and all participating study centres are obliged to support auditors and inspections of appropriate authorities, and to provide authorised persons access to original documents for this purpose.
All persons conducting audits are obliged to treat personal data and other data as confidential.

#### 6. Biometry

Statistical evaluation of the data will be performed by Dr. Florian Posch.
For the primary endpoint, the number of individuals with positive seroconversion to anti-SARS-CoV-2 spike protein at day 21-28 after the second vaccination measured by SARS-CoV-2 antigen-binding Ig assay will be assessed comparing immunocompromised patients to healthy controls.
**Null and alternative hypotheses:**
$H_0$: There is no statistical difference between the seroconversion rate of immunodeficient patients and healthy controls.
$H_1$: There is statistical difference between the seroconversion rate of immunodeficient patients and healthy controls.

##### *6. 1. Biometric design*

Group comparison, i.e. immunodeficient patients to healthy controls, will be performed for the primary as well as secondary endpoints according to the statistical methods described in 6.4.

Depending on the respective secondary endpoint, statistical analyses may be performed after visit 3, 4, 5 and/or 6.

##### *6. 2. Sampling plan*

To estimate the sample size for the primary endpoint, we considered levels of anti-SARS-CoV-2 IgG antibodies after 2nd vaccination. Therefore, we used nQuery, version 8.6.1 (Statistical Solutions Inc.), based on previous studies investigating the seroconversion of immunodeficient patients vaccinated against influenza (66). Mann–Whitney U test with a significance level of 0.01 and a power of 95% resulted in 175 subjects per group. Assuming 10% drop out rate, we calculated a sample size of 195 subjects per group.

##### *6. 3. Data registration and evaluation*

Statistical analysis of results will be done with IBM SPSS Statistics, version 26 or higher and R software (https://www.r-project.org/). Data quality will be assured by comparing random samples from processed data with the appropriate source data.

##### *6. 4. Statistical methods*

Primary endpoint:

The difference between healthy and immunodeficient individuals in the levels of anti-SARS-CoV-2 spike protein antibodies at day 21-28 after the second vaccination measured by SARS-CoV-2 antigen-binding Ig assay. Statistical significance will be calculated by comparison of immunocompromised patients to healthy controls via rank-sum tests. Patients developing SARS-CoV-2 infection or COVID-19 until primary endpoint evaluation will be omitted from the per-protocol analysis. P-values < 0.05 will be considered as statistically significant.

Secondary endpoints:

The levels of anti-SARS-CoV-2 spike protein antibodies at 6, 12 and 24 months after vaccination will be compared between immunocompromised patients and healthy controls by parametric or non-parametric tests (t-test, Mann-Whitney-U test, Kruskal-Wallis test), as appropriate. P-values < 0.05 will be considered as statistically significant.

Change of recombinant S protein-binding IgG levels after second vaccination to recombinant S protein-binding IgG levels after first vaccination will be compared between both time points using the one-sample t-test or Wilcoxon test, as appropriate. P-values < 0.05 will be considered as statistically significant.
Concentrations of secretory (saliva) and serum IgA as well as IgG and IgM after second vaccination will be treated as continuous variable and compared between immunocompromised patients and healthy controls by parametric or non-parametric tests (t-test, Mann-Whitney-U test, Kruskal-Wallis test), as appropriate. P-values < 0.05 will be considered as statistically significant.

IFNγ production of T cells after SARS-CoV-2 antigen exposure, measured by FACS and ELISpot, will be treated as continuous variable and compared between immunocompromised patients and healthy controls by parametric or non-parametric tests (t-test, Mann-Whitney-U test, Kruskal-Wallis test), as appropriate. P-values < 0.05 will be considered as statistically significant.

Baseline characteristics between immunocompromised patients and healthy controls will be compared with rank-sum tests, $\chi 2$-tests, and Fisher's exact tests, as appropriate. Associations of variables with high levels of anti-SARS-CoV-2 Ig will be studiesdin descriptive statistics. Although this is a hypothesis generating step, for further adequately power studies a correction for multiple testing will not be applied. P-values < 0.05 will be considered as statistically significant.

Neutralizing capacity of produced anti-SARS-CoV-2 IgG will be assessed in a descriptive manner.

For evaluation of dietary and/or body fat composition effects on anti-SARS-CoV-2 IgG levels, we will use Spearman's rank correlation or Pearson's correlation, as appropriate. P-values < 0.05 will be considered as statistically significant. Moreover, possible changes in body fat, ACE2 expression and dietary quality from visit 3 to 5 will be assessed in the studied groups via descriptive and explorative statistics.

Median follow-up time will be estimated with the reverse Kaplan-Meier method.

Risks of developing COVID-19, death-related-to-COVID-19 and death-from-other-causes will be calculated with competing risk cumulative incidence estimators Gray's tests, and Fine & Gray proportional subdistribution hazards models, respectively.

Missing data will not be replaced or estimated; only available data of participants will be considered in the analysis.

#### 7. Modification of the study protocol

The vote of the ethics committee applies solely to the information contained in the application; it does not include extensions or modifications of the research project undertaken at a later point in time. In case of any modification, an amendment of the study protocol signed by the Principal Investigator is required. Any modification of the study protocol must be attached as an amendment to all study protocols in circulation. The ethics committee must be informed of all modifications in the study protocol. In case of modifications in the study protocol that are not merely of a formal nature but contain changes pertinent to the study participants, a renewed vote of the ethics committee must be obtained. If applicable, the patients/probands must be informed in the information and consent form about changes in the terms and conditions of the study.

Substantial amendments must also be reported to the authorities (BASG). This is subject to non-prohibition in a period of 35 days. The ethics committee in charge must provide its statement within a period of 35 days.

#### 8. Ethical and legal matters

##### *8.1 Legal principles*

During the implementation of the trial, the (current versions of) following guidelines and laws must be followed in addition to the Declaration of Helsinki (such as):
• Current version of AMG (Austrian Act on Pharmaceutical Products)
• ICH-GCP guideline

• EU directives 2001/20/EC and 2005/28/EC, etc.

##### *8.2 Vote of the ethics committee;*

In accordance with § 40 AMG (Austrian Act on Pharmaceutical Products), the clinical trial may be started only after the competent ethics committee has issued its statement of approval and the appropriate authorities (BASG) have provided their non-prohibition/approval.

##### *8.3 Insurance*

During the clinical trial, a no-fault insurance (personal injury protection insurance in accordance with § 32 of the Austrian Act on Pharmaceutical Products) shall be concluded for the patient on behalf of the sponsor.
Exact contact data of the insurance company and the policy number is mentioned in the patient information form.

##### *8.4 Principal Investigator*

By signing the study protocol the Principal Investigator confirms that he/she has read and understood the study protocol, and will work in accordance with the protocol.
The Principal Investigator guarantees the confidentiality of all information.

##### *8.5 Storage and Data protection*

The registration, transfer, storage and evaluation of personal data in this clinical trial shall be subject to legal regulations (AMG - Austrian Act on Pharmaceutical Products - and data protection law). A prerequisite for this purpose is the participant's voluntary consent in the consent form, given prior to his/her participation in the clinical trial. The participants shall be informed of the following as part of the information about this clinical trial:

1. Data obtained in the course of this clinical trial shall be recorded on paper forms or electronic data storage devices, treated as strictly confidential, and passed on exclusively to the following persons without mentioning names (pseudonymised):

- the sponsor of the trial for scientific evaluation and assessment of adverse events.

2. If required for the inspection of the clinical trial, persons authorised by the sponsor and committed to secrecy (monitoring, auditing), authorised persons in national and foreign health authorities, and authorised persons of the competent ethics committee may inspect personal data at the study centre.

3. The participant shall be informed that he/she may terminate his/her participation in the clinical trial at any time without stating reasons and with no subsequent disadvantages. However, due to the legal obligation of documentation (AMG; Austrian Act on Pharmaceutical Products), authorised persons committed to secrecy may be permitted to inspect personal data for test purposes, for a period of time specified by law.

#### 9. Publication

Publication of data derived from this study requires the consent of the principal investigator and any co-principal investigator involved. Publications with the main focus on humoral immunity shall be coordinated by Ivo Steinmetz. Publications with the main focus on T cell immunity, CVID or rituximab shall be coordinated by Martin Stradner. Publications with the main focus on vaccination in hematologic diseases shall be coordinated by Hildegard Greinix. The coordinating authors will appoint junior authors. Coordinating and junior authors are responsible for the preparation of the manuscripts. Authorship will be granted according the Uniform Requirements for Manuscripts Submitted to Biomedical Journals.

### 11. Signatures

#### *11.1 Sponsor*

By signing this document I confirm that the trial shall be performed in accordance with ICH-GCP, the Declaration of Helsinki, national laws, and the current study protocol.

**Sponsor or his representative**

STRADNER MARTIN 9.04.2021 [signature]

**Name, First name (in block letters) Date, Signature**

##### 11.2 Principal Investigator

I confirm herewith that I have read and understood the present study protocol, and acknowledge all parts of it. I promise to ensure that the persons introduced in the trial at my centre shall be treated, observed and documented in accordance with the terms and conditions of this study protocol.

STRADNER MARTIN

**Name, First name (in block letters)**

9.4.2021 [signature]

**Date, Signature**

#### Appendix

##### *Product characteristics (in German)*

Comirnaty: https://www.ema.europa.eu/en/documents/product-information/comirnaty-epar-product-information_de.pdf

Moderna Covid-19 vaccine: https://www.ema.europa.eu/en/documents/product-information/covid-19-vaccine-moderna-epar-product-information_de.pdf

AstraZeneca COVID-19 vaccine: https://www.ema.europa.eu/en/documents/product-information/covid-19-vaccine-astrazeneca-epar-product-information_de.pdf
